# Rural-to-Urban Migrant Worker Mobility Shaped Measles Epidemics in China

**DOI:** 10.1101/2025.06.21.25330021

**Authors:** Peihua Wang, Xianwen Wang, Wenyi Zhang, Yong Wang, Sen Pei, Xiao-Ke Xu, Wan Yang

## Abstract

Despite sustained high routine childhood vaccination coverage, measles outbreaks have persisted across Provincial-Level Administrative Divisions (PLADs) in China. Epidemiological evidence suggests that migrant workers substantially contribute to these outbreaks. In this study, we investigated the role of inter-PLAD rural-to-urban migrant workers—a migrant worker group originating from less developed rural regions with potentially lower vaccination coverage—in driving measles epidemics in China from 2005 to 2014. We developed a networked metapopulation Susceptible–Exposed–Infectious– Recovered model incorporating detailed migrant worker mobility and traveler mobility patterns. By simulating measles transmission dynamics within migrant worker subpopulations, we identified key epidemiological connections between origin and host PLADs. In northern China, migrant workers from Hebei and Shandong were the key contributors to outbreaks in two northern host PLADs, Beijing and Tianjin. In southern China, migrant workers from Anhui and Sichuan were the key contributors across multiple southern host PLADs. Counterfactual modeling suggests that measles epidemics in host PLADs were driven by susceptibility replenishment from under-vaccinated migrant workers during the Chinese New Year migration periods. Moreover, epidemics in origin PLADs might have been synchronized and facilitated by case importation from migrant workers returning from endemic host PLADs, and the strength of this seeding effect depended on the volume of migrant worker flows. Traveler mobility showed minimal impact on measles epidemics. Counterfactual modeling of pre-migration vaccination showed a 50.9% incidence reduction nationally, with significant reduction in host PLADs, and in turn in origin PLADs due to weakened seeding effect. Our findings provide mechanistic insights into the epidemiological role of rural-to-urban migrant workers in measles epidemics, which could support targeted vaccination strategies for improved measles control in China and regions with similar migration dynamics.

## Introduction

Measles is a highly infectious disease with an estimated basic reproductive number (*R_0_*; i.e., average number of secondary infections caused by one infected individual in a fully susceptible population) of 12 to 18 (1, 2). Vaccination in children has been the most effective public health intervention, and models predict that at least 95% vaccination coverage is needed for measles elimination given its *R_0_* (3, 4). In China, routine measles vaccination was included in the national Expanded Program on Immunization (EPI) in 1978, and a two-dose schedule was introduced in 1986 (5). However, despite reported routine childhood vaccination coverage exceeding 95% since 2005 in Provincial-Level Administrative Divisions (PLADs) such as Beijing (6), Tianjin (7), and other economically developed PLADs (5), annual measles outbreaks persisted through 2010. The nationwide Supplementary Immunization Activity (SIA) in 2010, targeting children aged 1–14 years, temporally curbed transmission, but outbreaks rebounded between 2013 and 2016. These immunization programs substantially reduced measles transmission among children, which was likely the primary driver of outbreaks when vaccination coverage was not sufficiently high (8). Nevertheless, epidemiological studies in China have documented a shift towards increased prevalence among adults, particularly migrant workers originating from less developed regions (9–11). These migrant workers generally have higher susceptibility due to limited healthcare accessibility and consequently lower vaccination coverage in their hometowns (12, 13). Modeling studies in China (14, 15) further support these epidemiological findings, suggesting that in addition to climate factors modulating measles transmission, migrant worker mobility contributes to measles outbreaks by replenishing susceptible population in host PLADs.

Migrant workers constitute a substantial portion of the Chinese population (∼19.6% of total population in 2010 (16)) and primarily comprise two types: rural-to-urban migrant workers and urban-origin white-collar migrant workers. Rural-to-urban migrant workers originate from rural areas (i.e., holding rural household registrations within the Chinese household registration system, which officially identifies individuals as permanent residents of specific locations and determines their access to local social welfare, healthcare, and educational benefits (17)). They migrate to urban areas and are predominantly employed in manufacturing, construction, wholesale, and retail sectors (18). In contrast, urban-origin white-collar migrant workers typically have higher education levels and are employed in professional and managerial positions within urban centers (19). Due to lower economic development and limited healthcare accessibility in their rural hometowns, combined with limited healthcare benefits associated with rural household registrations, rural-to-urban migrant workers likely have lower measles vaccination coverage compared to white-collar migrant workers, and thus may play a crucial role in measles epidemics.

From 2005 to 2014, inter-PLAD rural-to-urban migrant workers (hereafter referred to as migrant workers) averaged ∼75 million annually, accounting for 5.7% of the national population in China (20). This population typically migrated to economically developed PLADs following the Chinese New Year (CNY) holiday to seek employment. Such large-scale seasonal migration is analogous to migration driven by agricultural cycles, which has been shown to shape measles epidemics in Africa (21, 22), and could rapidly alter the contact patterns and population immunological profiles in host PLADs. Notably, this migrant worker influx temporally coincided with epidemic peaks around April (15). Given the magnitude and timing of worker migration, detailed mechanistic modeling of rural-to-urban migrant worker mobility is needed to quantitatively assess its impact on measles epidemics.

In this study, we investigated the role of inter-PLAD rural-to-urban migrant workers in driving measles epidemics in host PLADs from 2005 to 2014. We developed a networked metapopulation Susceptible–Exposed–Infectious–Recovered (SEIR) model that incorporates detailed migrant worker mobility data. Using this model, we simulated transmission dynamics within migrant worker subpopulations, identified key epidemiological connections between origin and host PLADs, and quantified the impacts of migrant worker mobility and population immunological profiles on measles epidemics. Our results provide mechanistic insights into the epidemiological role of rural-to-urban migrant workers in measles epidemics, which could inform targeted public health interventions to control measles transmission in China.

## Results

### Mobility patterns and their associations with measles incidence

Time series of inter-PLAD traveler network (general-purpose travel, including tourism, business trips, and white-collar worker migration) from 2005 to 2014 was used to compute network characteristics of PLADs. PLADs such as Beijing, Tianjin, Shanghai, Jiangsu, Zhejiang, Fujian, and Guangdong showed increased population outflows before the CNY holidays (Fig. S1) and inflows afterward (Fig. 1a). These PLADs had the highest per-capita Gross Regional Product (GRP) (20) and had the largest proportions of inter-PLAD rural-to-urban migrant workers (≥8.6%; Fig. S2), thus serving as migrant worker host PLADs. Conversely, less developed PLADs, home to most migrant workers, showed reversed mobility flows. These seasonal mobility patterns primarily resulted from migrant workers returning to their origin PLADs before the CNY holidays for family reunions and to host PLADs for employment afterward.

**Fig. 1.**
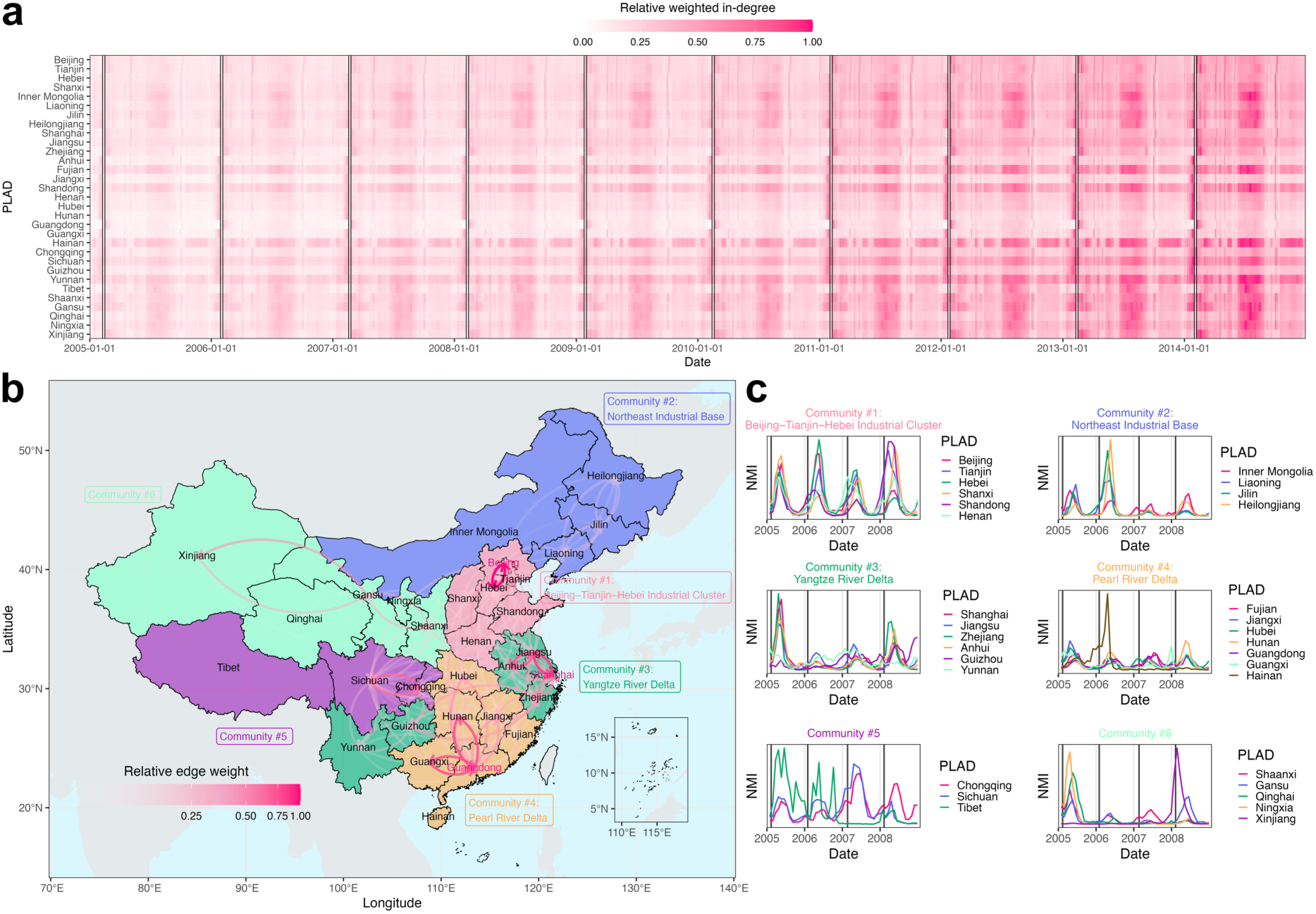
(**a**) Relative weighted in-degree of the inter-PLAD traveler network from 2005 to 2014. Boxes indicate CNY holiday periods. (**b**) Infomap community detection based on aggregated mobility patterns six weeks before and after the CNY holidays. Directed edges indicate mobility flows, and PLADs highlighted in red indicate transportation hubs. (**c**) Normalized monthly incidence (NMI) of measles in PLADs within each community from 2005 to 2008, a period characterized by regular annual outbreaks before the nationwide SIA in 2010. Vertical lines indicate the first day of each CNY holiday.

We constructed an aggregated traveler network by summing traveler volumes (edge weights) between each PLAD pair (node pair) over the migration periods (six weeks before and after the CNY holidays) from the original time-series traveler network. Infomap community detection (23) was then applied to partition the network by minimizing the description length of random walks, resulting in six communities characterized by dense internal traveler flows (Fig. 1b). Four of these overlapped with key economic zones in China: the Beijing–Tianjin–Hebei Industrial Cluster (community #1) and the Northeast Industrial Base (historically a heavy industry center; community #2) in the north, the Yangtze River Delta (community #3) in the east, and the Pearl River Delta (community #4) in the south. Beijing (in community #1), Shanghai (in community #3), and Guangdong (in community #4) were major transportation hubs with the highest PageRank centralities, collectively accounting for 47.0% of all flows during the CNY migration period.

During the study period, measles outbreaks occurred with varying patterns across PLADs. PLADs in the Beijing–Tianjin–Hebei Industrial Cluster and Pearl River Delta showed annual measles outbreaks of similar magnitude (excluding certain PLADs with highly irregular outbreaks, Fig. 1c). In contrast, PLADs in the Northeast Industrial Base and Yangtze River Delta also showed annual outbreaks but of differing magnitudes (Fig. 1c), whereas PLADs in western China showed less synchronized outbreaks (Fig. 1c). Nonetheless, measles incidence in PLADs within the same communities showed higher correlations than that between communities (Mantel test, *r* = 0.10, *P* = 0.014).

### Modeled population dynamics and measles epidemic dynamics

The networked metapopulation SEIR model integrated two mobility networks: the traveler network (representing general-purpose travel) operating throughout the year, and the migrant worker network (representing rural-to-urban worker migration specifically) operating only during the migration periods (six weeks before and after the CNY holidays). The model defined seven migrant worker host PLADs (Beijing, Tianjin, Shanghai, Jiangsu, Zhejiang, Fujian, and Guangdong) based on their having the largest proportions of migrant workers and the highest per-capita GRP, and 18 migrant worker origin PLADs based on their population sizes within host PLADs (see Fig. S2 for detailed host–origin PLAD pairs). Two host PLADs, Jiangsu and Zhejiang, also served as origins for migrant workers. The remaining eight PLADs, including western PLADs (Yunnan, Tibet, and the five PLADs in community #6; Fig. 1b) and the island PLAD (Hainan), were neither host nor origin PLADs.

In host PLADs, such as Jiangsu (Fig. 2a) and Beijing (Fig. 2b), migrant worker populations (indicated as “host_origin”) tended to decrease during the 6-week pre-CNY period as migrant workers returned to the origin PLADs for family reunions, and increase during the 6-week post-CNY period as they moved into host PLADs for employment.

**Fig. 2.**
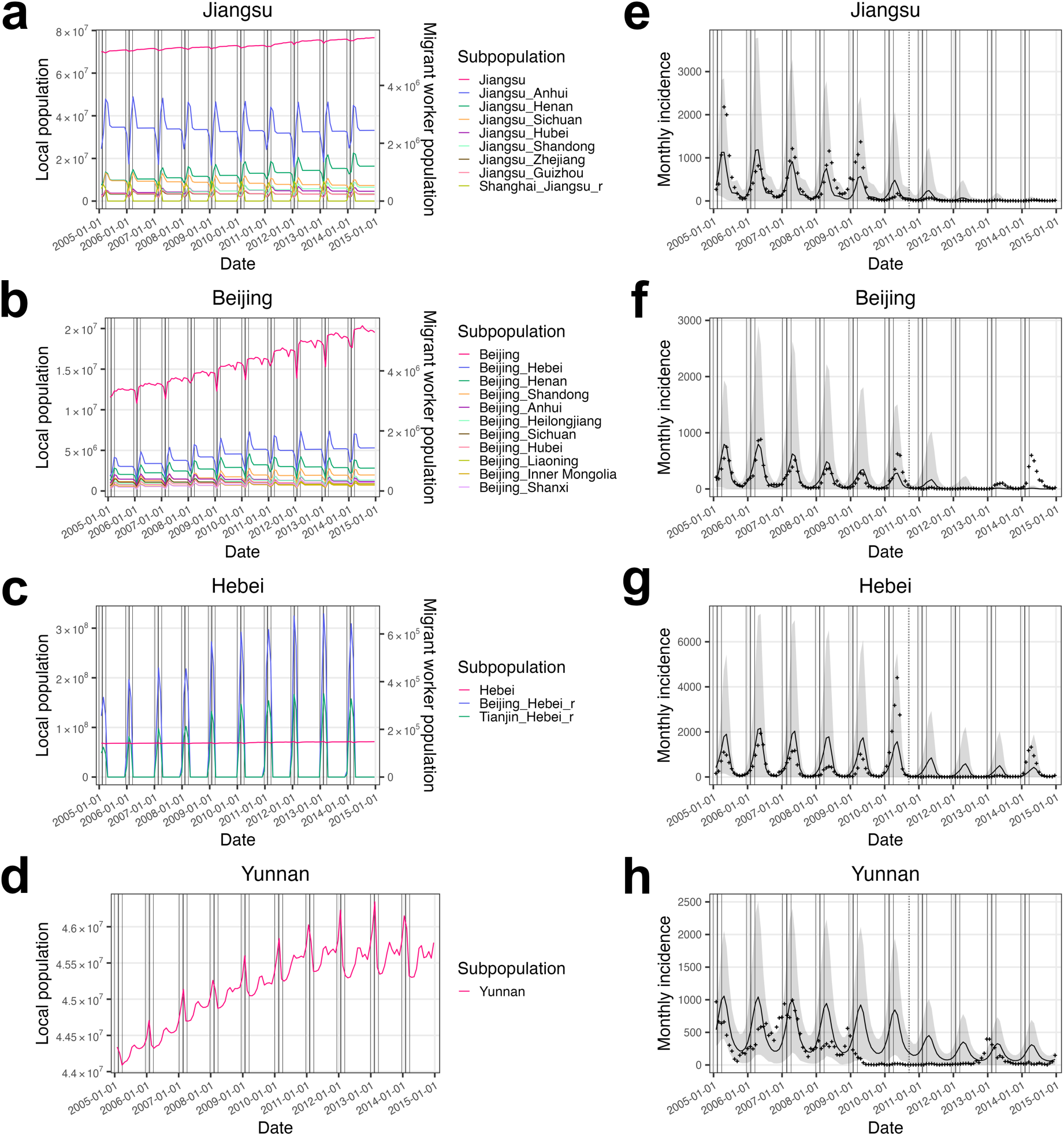
Population dynamics of local and migrant worker subpopulations illustrated using examples from (**a**) Jiangsu (both a host and an origin PLAD), (**b**) Beijing (a host PLAD), (**c**) Hebei (an origin PLAD), and (**d**) Yunan (neither a host nor an origin PLAD). Subpopulation labeled “PLAD A_PLAD B” indicate a migrant worker subpopulation with host PLAD A and origin PLAD B. Subpopulation labeled “PLAD A_PLAD B_r” indicate a returned migrant worker subpopulation with host PLAD A and origin PLAD B. Migrant worker subpopulations in the legend are ordered by their population sizes from the largest at the top to the smallest at the bottom. Vertical solid lines from left to right indicate the starts of pre-CNY, CNY, and post-CNY periods, and the end of post-CNY period for each year. Simulated measles incidence from the calibrated model ensemble for (**e**) Jiangsu, (**f**) Beijing, (**g**) Hebei, and (**h**) Yunnan, compared with observed incidence (crosses). Lines indicate median estimates, and shaded area indicates 95% confidence interval. The vertical dashed line indicates the nationwide SIA in 2010.

Correspondingly, in origin PLADs, such as Jiangsu (Fig. 2a) and Hebei (Fig. 2c), migrant worker populations returning from host PLADs (indicated as “host_origin_r”, e.g., “Beijing_Hebei_r” in Hebei) showed dynamics opposite to their counterparts in host PLADs (e.g., “Beijing_Hebei” in Beijing). In the model, the migrant worker flow during the post-CNY period included both recurring and new migrant workers. Their populations in host PLADs peaked at the end of the post-CNY migration period and tended to decrease afterward (Figs. 2a and b), as migrant workers who were unable to secure employment returned to their origin PLADs. PLADs that were neither host nor origin PLADs, such as Yunnan (Fig. 2d), included only local populations. The dynamics of local populations in all PLADs were driven by mobilities including tourism, business trips, and white-collar worker migration (note that white-collar migrant workers were treated as part of the local population in the model because their population susceptibility was comparable to that of local residents), in addition to births and deaths.

Incorporating the above population dynamics, we calibrated the model using monthly measles incidence data from 2005 to 2008 (a period characterized by regular annual outbreaks in China and before the nationwide SIA in 2010) for all PLADs (Figs. 2e–h and S3–6). The calibrated model showed reasonable overall model fit (mean correlation coefficient across PLADs = 0.566, mean peak time difference = 1.08 months; see details of the metrics in Table S1). It captured regular annual outbreaks, and larger observed outbreaks generally fell within the 95% confidence intervals (e.g., Hebei, Fig. 2g). Host PLADs and origin PLADs connected through the migrant worker network showed better model fits than the other disconnected western and island PLADs (comparison based on relative root mean square error, Mann–Whitney *U* test, *z* = 0.41, *P* = 0.023; Table S1). These disconnected PLADs experienced irregular epidemic dynamics characterized by large outbreaks followed by epidemic fadeout (Fig. 1c), suggesting influences other than mobility, such as higher birth rates (Mann–Whitney *U* test, *z* = 0.57, *P* = 0.001) and variability in vaccination coverage.

### Key migrant worker subpopulations contributing to measles outbreaks in host PLADs

The calibrated model ensemble simulated measles incidence for each subpopulation within PLADs (Figs. 3a–d). By ranking incidence within each host PLAD, we identified top-ranked migrant worker subpopulations that collectively accounted for >50% of total incidence from migrant workers (17 out of 52 in total, red arrows indicate the subpopulations contributing the highest incidence, and blue arrows indicate additional top-ranked subpopulations, Fig. 3e). In northern China, Hebei and Shandong were key contributors to outbreaks in both northern host PLADs, namely Beijing and Tianjin (connections aligned with Infomap community #1, corresponding to the Beijing–Tianjin–Hebei Industrial Cluster, Fig. 1b). In southern China, Anhui and Sichuan were the key contributors to outbreaks across multiple southern host PLADs. Specifically, Anhui was the primary contributor in Jiangsu, Shanghai, and Zhejiang (Infomap community #3, corresponding to the Yangtze River Delta, Fig. 1b). Sichuan was the key contributor across all southern host PLADs. It was the only western key PLAD, and it formed long-distance epidemiological connections to host PLADs.

**Fig. 3.**
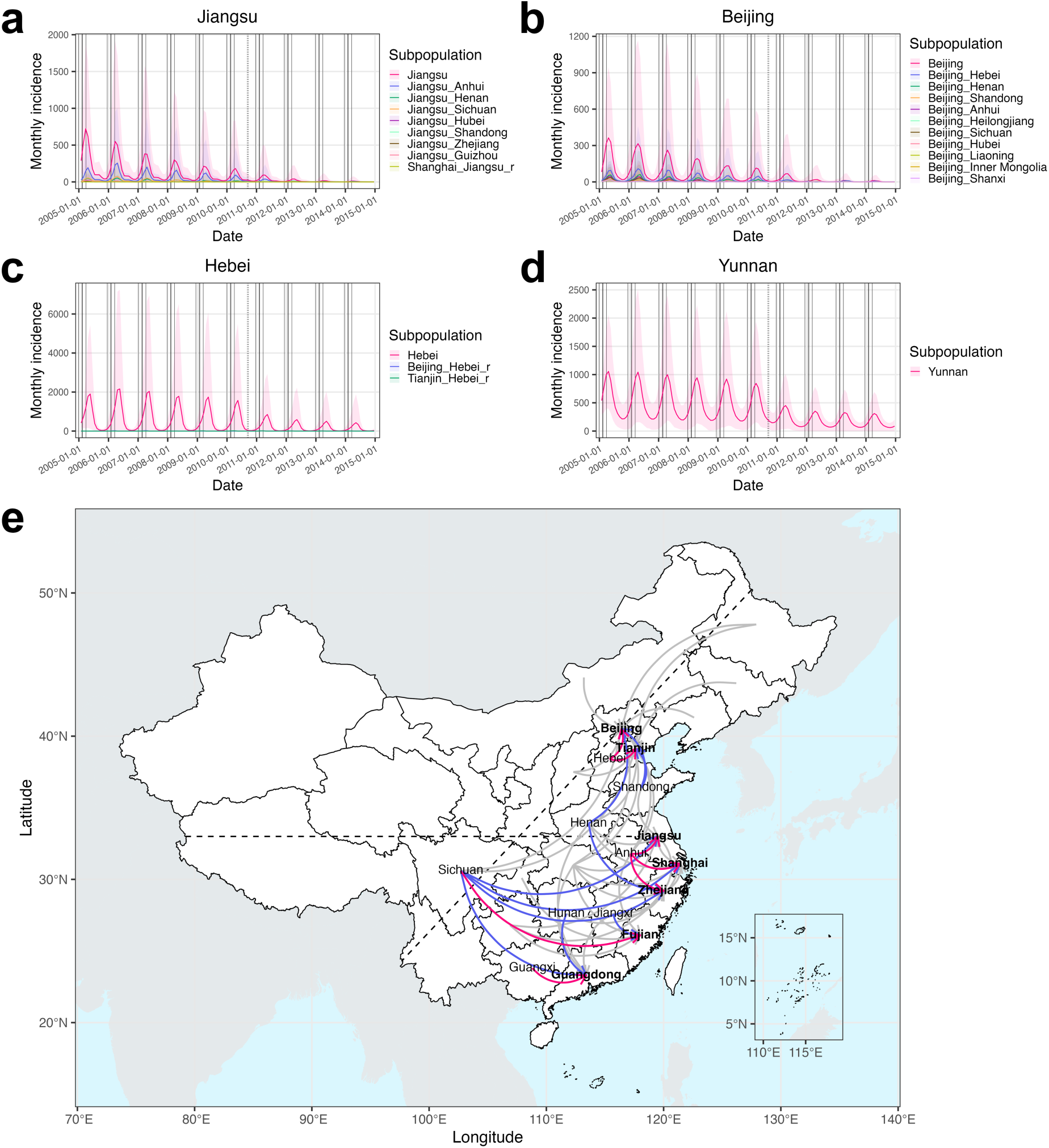
Simulated measles incidence from the calibrated model ensemble for each subpopulation, illustrated using examples from (**a**) Jiangsu, (**b**) Beijing, (**c**) Hebei, and (**d**) Yunnan. (**e**) Key epidemiological connections among PLADs. Arrows indicate migrant worker subpopulations moving from origin (tail) to host (head) PLADs. Arrows highlighted in red indicate migrant worker subpopulations that contributed the highest measles incidence among all migrant worker subpopulations in their corresponding host PLADs. Arrows highlighted in blue indicate additional migrant worker subpopulations that were among the top-ranked contributors to incidence in their corresponding host PLADs. Host PLADs are indicated in bold font. The horizontal dashed line approximates the Qin Mountains–Huai River reference line at ∼33°N, which divides China into northern and southern regions, and the tiled dashed line indicates the Heihe–Tengchong reference line, which divides China into western and eastern regions.

Additionally, Guangxi and Hunan were key contributors in Guangdong (Infomap community #4, corresponding to the Pearl River Delta, Fig. 1b), and Jiangxi was a key contributor in Fujian (Infomap community #4, Fig. 1b). Henan contributed substantially in both northern (Beijing) and southern (Zhejiang) host PLADs.

### Impacts of mobility and population immunological profiles assessed through counterfactual modeling

We tested the impacts of traveler mobility (spatial spread) and migrant worker mobility (case importation and susceptibility replenishment) on measles epidemics by using counterfactual modeling and comparing the simulated cumulative incidence with that of the baseline scenario from the calibrated model ensemble. In the first analysis, we removed traveler mobility (“no travelers” scenario). The results showed minimal impact of non-migration-related general travels on measles epidemics (Tables 1, S2 and S3; Figs. 4a–d).

**Fig. 4.**
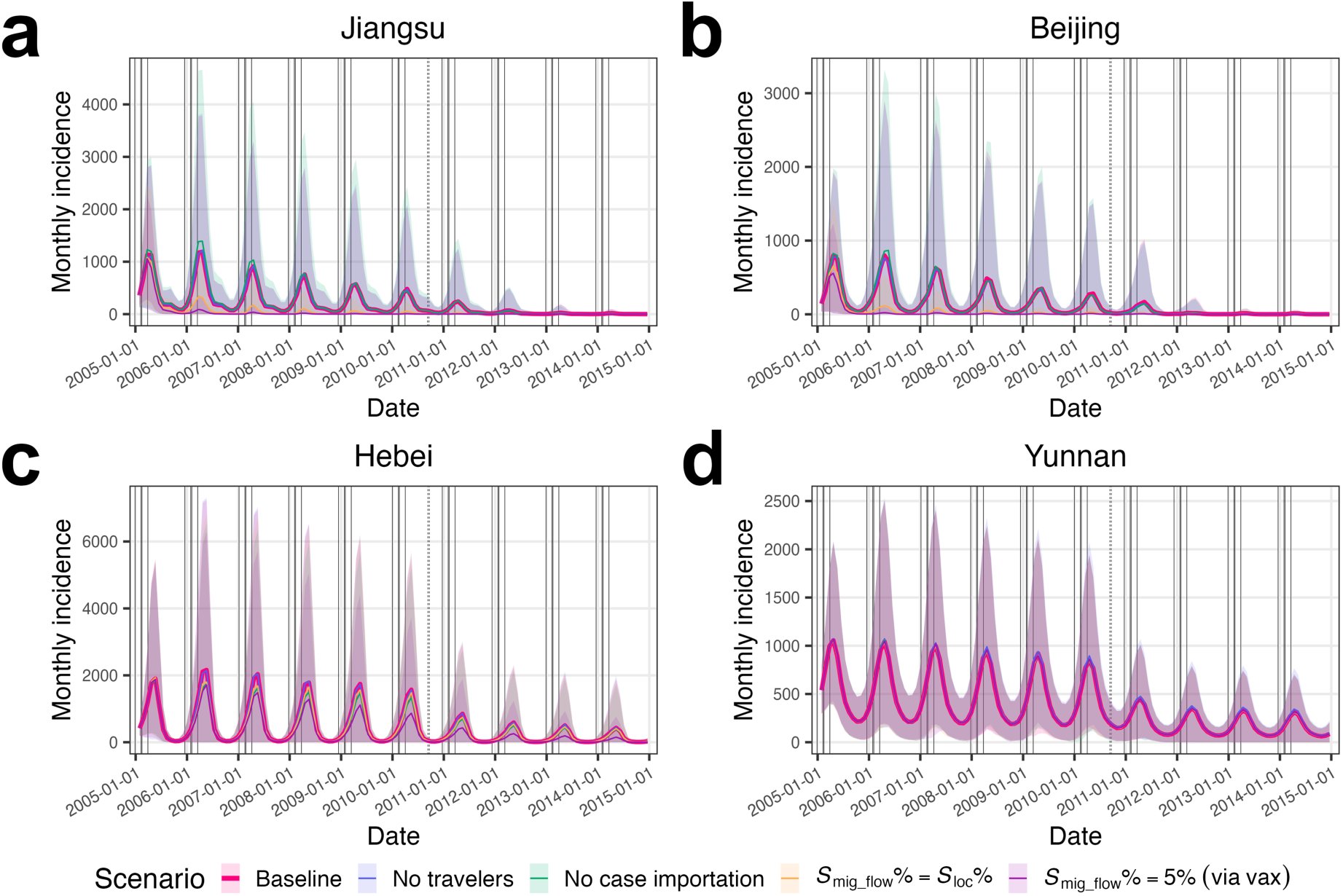
Simulated measles incidence under the baseline scenario compared with that under counterfactual scenarios in (**a**) Jiangsu, (**b**) Beijing, (**c**) Hebei, and (**d**) Yunnan. Counterfactual scenarios include no travelers, no case importation, matching population susceptibility between incoming migrant workers and local populations in host PLADs, and pre-migration vaccination for migrant workers.

The second analysis, removing the movement of exposed and infectious migrant workers (“no case importation” scenario), estimated a statistically significant 15.4% reduction in national measles incidence, largely due to reductions in origin PLADs (Tables 1 and S2). Specifically, this scenario showed no significant change in incidence in host PLADs (see Jiangsu in Fig. 4a and Beijing in Fig. 4b as examples), except for Shanghai, where incidence decreased (Table 1). This decrease was likely due to the already low incidence rates in Shanghai starting from 2006 (Fig. 1c; average incidence rate = 4.3/100,000 population/year during 2006–2007, the lowest among host PLADs) and the limited susceptibility replenishment from connections to Jiangsu and Zhejiang, origin PLADs that had lower population susceptibilities (before the nationwide SIA in 2020, *S*_Jiangsu_% = 6.2%, *S*_Zhejiang_% = 6.0%, both lower than *S*_Shanghai_% = 6.3%; Fig. S7a). Origin PLADs showed varying degrees of incidence reduction (Table S2; see Hebei in Fig. 4c as an example). Origin PLADs with larger migrant worker proportions had greater incidence reductions (Spearman’s rank correlation test, ρ = 0.85, *P* = 3.1×10^-5^), with 7 of 16 origin PLADs (excluding Jiangsu and Zhejiang) showing significant reductions. PLADs that were neither hosts nor origins were minimally impacted (Table S3; see Yunnan in Fig. 4d as an example). Taken together, these results suggest that migrant workers might have facilitated the introduction of measles from their host PLADs to their origin PLADs.

**Table 1.** Relative differences in cumulative measles incidence between counterfactual scenarios and the baseline in host PLADs. Values indicate mean estimates with 95% confidence intervals (calculated using the bootstrap method).

| China or host PLAD | Counterfactual scenario |  |  |  |
| --- | --- | --- | --- | --- |
|  | No travelers | No case importation | Matching population susceptibility | Pre-migration vaccination |
| China | -0.3%<br>(-6.6%, 6.4%) | -15.4%<br>(-21.9%, -8.5%) | -35.8%<br>(-40.3%, -31.1%) | -50.9%<br>(-54.2%, -47.2%) |
| Beijing | -0.7%<br>(-19.1%, 19.7%) | 6.4%<br>(-15.2%, 31.9%) | -69.6%<br>(-74.3%, -65.0%) | -79.1%<br>(-82.2%, -76.0%) |
| Tianjin | 0.6% | 0.0% | -73.9% | -83.9% |
|  | (-22.9%, 26.8%) | (-22.5%, 25.6%) | (-78.7%, -68.6%) | (-86.8%, -80.8%) |
| Shanghai | -1.7%<br>(-21.5%, 22.6%) | -22.7%<br>(-41.1%, -0.2%) | -51.5%<br>(-59.9%, -42.2%) | -73.7%<br>(-78.1%, -69.0%) |
| Jiangsu | -0.6%<br>(-20.8%, 23.1%) | 3.8%<br>(-18.4%, 30.6%) | -74.5%<br>(-78.8%, -70.3%) | -84.0%<br>(-86.6%, -81.3%) |
| Zhejiang | 0.7%<br>(-18.1%, 23.1%) | 10.6%<br>(-12.4%, 37.9%) | -72.6%<br>(-76.6%, -67.6%) | -82.3%<br>(-84.9%, -79.4%) |
| Fujian | 1.8%<br>(-27.2%, 37.0%) | 1.8%<br>(-28.3%, 39.3%) | -77.4%<br>(-82.8%, -72.2%) | -84.8%<br>(-88.1%, -81.1%) |
| Guangdong | -0.2%<br>(-16.5%, 18.5%) | 13.8%<br>(-8.6%, 38.9%) | -69.9%<br>(-75.3%, -64.1%) | -89.2%<br>(-91.0%, -87.0%) |

Our calibrated model estimated higher population susceptibility in origin PLADs than that in host PLADs during 2005–2014 (before the nationwide SIA in 2010: Wilcoxon rank-sum test, *P* = 3.8×10^-7^, average *S*_host_% = 6.2%, average *S*_origin_% = 7.4%, Fig. S7a; after the nationwide SIA: *P* = 3.8×10^-7^, average *S*_host_% = 6.1%, average *S*_origin_% = 7.0% Fig. S7b). To test the impact of susceptibility replenishment from migrant workers, we conducted a third analysis assuming equal population susceptibility between incoming migrant workers and local populations in host PLADs (“matching population susceptibility” scenario). This scenario estimated a significant 35.8% national incidence reduction and significant reductions across host PLADs (Table 1). Shanghai showed a smaller reduction compared with other host PLADs, likely due to its connections with Jiangsu and Zhejiang, origin PLADs with lower susceptibility. The impact on host PLADs became evident one year into the simulation (see Jiangsu in Fig. 4a and Beijing in Fig. 4b as examples), as shown by epidemic fadeouts from summer through early winter in 2005, and the subsequent 72.5% average incidence reduction compared with baseline before migrant workers began arriving in host PLADs. In origin PLADs, greater incidence reductions were associated with larger migrant worker proportions (Spearman’s rank correlation test, ρ = 0.64, *P* = 0.008), with one origin PLAD showing significant reduction (Table S2). PLADs that were neither hosts nor origins were minimally impacted (Table S3). These results suggest that susceptibility replenishment from under-vaccinated migrant workers significantly contributed to measles outbreaks in host PLADs.

Lastly, in the fourth analysis, we tested a potential intervention of vaccinating migrant workers prior to their departure (“pre-migration vaccination” scenario). Assuming full vaccination coverage and 95% vaccine effectiveness, this scenario estimated a significant 50.9% national incidence reduction and significant reductions across host PLADs (Table 1). Similar to the two previous scenarios, origin PLADs with larger migrant worker proportions showed greater incidence reductions (Spearman’s rank correlation test, ρ = 0.95, *P* = 2.7×10^-8^), although this effect was more pronounced, with 10 out of 16 origin PLADs showing significant reductions (Table S2). As with the other scenarios, minimal impacts were seen in PLADs that were neither hosts nor origins (Table S3).

## Discussion

In this study, we investigated the role of inter-PLAD rural-to-urban migrant workers in driving measles epidemics in China from 2005 to 2014. We developed a metapopulation SEIR model integrated with a migrant worker network and a traveler network to simulate measles transmission dynamics in both migrant worker and local subpopulations. Using this model, we identified key migrant worker subpopulations that contributed substantially to measles outbreaks in host PLADs despite sustained high vaccination coverage exceeding 95% in these host PLADs, and quantified the impacts of migrant worker mobility on measles epidemics through case importation and susceptibility replenishment.

The model identified key epidemiological connections between origin and host PLADs. In northern China, the model suggests that migrant workers from Hebei and Shandong were key contributors to outbreaks in Beijing and Tianjin. These PLADs were grouped together in the Infomap community #1 (Fig. 1b) corresponding to the Beijing– Tianjin–Hebei Industrial Cluster. In southern China, the inferred connections were more complex. Anhui and Sichuan were identified as key origin PLADs contributing to outbreaks across multiple southern host PLADs. Specifically, migrant workers from Anhui contributed substantially to outbreaks in Jiangsu, Shanghai, and Zhejiang, corresponding to the Yangtze River Delta (Infomap community #3, Fig. 1b), and those from Sichuan contributed substantially in all southern host PLADs. In contrast to other key epidemiological connections characterized by geographic contiguity, those originating from Sichuan represented long-distance epidemiological connections between the western origin PLAD and eastern host PLADs. Additionally, Henan contributed substantially to outbreaks in both northern (Beijing) and southern (Zhejiang) host PLADs. While previous epidemiological studies typically reported aggregated incidence among migrant workers (9–11), our results provide detailed origin-host epidemiological connections that could support targeted vaccination strategies for measles control.

Counterfactual simulation removing traveler mobility (“no travelers” scenario) showed minimal impacts on measles epidemics (national incidence reduced by 0.3% compared with the baseline, Table 1). Given low measles incidence across PLADs during the study period (average annual incidence rate: 7.1/100,000 population/year from 2005 to 2010 before the nationwide SIA, and 1.8/100,000 population/year from 2011 to 2014), traveler-related seeding effect was likely limited. Additionally, because travelers typically remained in destination PLADs only briefly, their mobility had little influence on local population susceptibility. This finding contrasts with other respiratory infectious diseases, such as influenza (24–26) and SARS-CoV-2 (26, 27), for which travelers or commuters facilitated broader geographic spread, potentially due to stronger seeding effect or higher population susceptibility.

Counterfactual simulation removing case importation through migrant worker mobility (“no case importation” scenario) resulted in a 15.4% reduction in national measles incidence (Table 1), primarily in origin PLADs. The magnitude of incidence reduction was positively correlated with the proportion of migrant workers in origin PLADs, suggesting that the strength of this seeding effect depended on the volume of migrant worker flows returning to their origin PLADs during the pre-CNY periods. These results emphasize the importance of controlling measles transmission in host PLADs, as endemic transmission therein could seed outbreaks in migrant worker origin PLADs. Effective control in host PLADs would therefore reduce measles incidence locally and in the connected origin PLADs.

Our model estimated higher population susceptibility in origin PLADs compared to host PLADs. To test the impact of susceptibility replenishment from migrant workers, we simulated a counterfactual scenario in which incoming migrant workers had susceptibility equal to that of local subpopulations in host PLADs (“matching population susceptibility” scenario). This scenario resulted in a 35.8% reduction in national incidence (Table 1), with significant reductions in host PLADs and varying reductions in origin PLADs. This result suggests that susceptibility replenishment from under-vaccinated migrant workers significantly contributed to measles outbreaks in host PLADs. Additionally, the reduced incidence in host PLADs led to further reductions in origin PLADs by weakening the seeding effect, and the degree of which was positively correlated with the proportion of migrant workers in origin PLADs.

The results from the counterfactual scenario, in which population susceptibility was matched between incoming migrant workers and local subpopulations in host PLADs, suggest that operational intervention strategies such as pre-migration vaccination could effectively control measles transmission in host PLADs and in turn reduce incidence in origin PLADs. Our counterfactual simulation of vaccinating migrant workers before their departure (“pre-migration vaccination” scenario), assuming full coverage and 95% vaccine effectiveness, resulted in the largest reduction in incidence nationwide (Table 1). Significant reductions occurred in all host PLADs and in 10 of the 16 origin PLADs (origin PLADs excluding Jiangsu and Zhejiang), with larger reductions in origin PLADs having higher migrant worker proportions. Beijing has implemented migrant worker-targeted SIAs since 2005 (e.g., 0.8 million vaccinated among 5.7 million migrant workers in 2009) (28, 29), which helped reduce, though not eliminate, measles transmission. Here, our results suggest that achieving higher vaccination coverage and implementing synchronized vaccination campaigns targeting migrant workers prior to their departure to host PLADs may more effectively control endemic transmission in host PLADs, origin PLADs, and throughout China.

Our study has several limitations. First, we only modeled measles transmission at the PLAD level, so we could not capture inhomogeneous mixing and migrant worker mobility at finer spatial resolutions. Thus, we were unable to simulate granular, localized outbreaks or the irregular outbreaks observed in PLADs that were neither hosts nor origins. Second, we did not incorporate age structure, which may oversimplify age-dependent immunological profiles. Third, although we attempted to model the mobility of transient migrant workers who failed to secure employment, in addition to long-term migrant workers, we did not fully capture the magnitude and complexity of their mobility patterns due to limited data. Despite these limitations, our study identified detailed epidemiological connections between PLADs and quantified the impact of inter-PLAD rural-to-urban migrant worker mobility on measles epidemics. In host PLADs with high routine childhood vaccination coverage, outbreaks persisted primarily due to susceptibility replenishment from under-vaccinated migrant workers. In origin PLADs, measles epidemics might have been synchronized and facilitated by case importation from migrant workers returning from endemic host PLADs. These findings offer mechanistic insights into how migrant worker mobility shaped measles epidemic dynamics, which could inform public health interventions for measles control in China.

## Methods and materials

### Study data

Monthly measles incidence data for each PLAD from 2005 to 2014 were sourced from the Data-center of China Public Health Science (30). City-level daily mobility data from February 3, 2015, to February 2, 2019, were sourced from Tencent mobile device location service (travel purposes unavailable). The dataset included 32.6 billion travel records from 363 of 368 cities at the prefecture, sub-provincial, and provincial levels in China.

Demographic data, including birth and death rates, the sizes of total populations, and per-capita GRP were obtained from the National Bureau of Statistics of China (20).

Immunization rates were estimated as described previously (15). Details on the estimations of inter-PLAD rural-to-urban migrant worker population sizes are provided in the Supplementary Text “Estimation of inter-PLAD rural-to-urban migrant worker population sizes”.

### Construction of inter-PLAD traveler network and migrant worker network during 2005–2014

We constructed the inter-PLAD traveler network and migrant worker network from 2005 to 2014 based on the 2015–2019 mobility data. To temporally align mobility patterns from 2015–2019 to the 2005–2014 study period for network construction, we aggregated the original mobility data based on national holidays and inter-holiday periods. Specifically, edge weights (mobility flow volumes) between the same node pairs (departure– arrival city pairs) were averaged for corresponding days within each holiday or inter-holiday period. To match the spatial resolution of our analyses at the PLAD level, we further aggregated these city-level data by summing edge weights for the same departure–arrival PLAD pairs.

To construct the traveler network representing general-purpose travel, we assumed that traveler mobility patterns during 2005–2014 followed the aggregated holiday and inter-holiday mobility patterns. Specifically, we divided the 2005–2014 study period into national holidays and inter-holiday periods, and assigned flow volumes from the aggregated mobility data to corresponding days within each period. These daily flow volumes were then scaled to match reported annual travel volumes (20). To ensure flow mass balance (i.e., balance between cumulative inflow and outflow volumes through each PLAD over the 10-year period), traveler volumes were adjusted using iterative proportional fitting (31). The resulting scaling factors ranged from 0.92 to 1.08.

To construct the migrant worker network representing rural-to-urban worker migration specifically, we assumed that migrant worker mobility during 2005–2014 was concentrated within the migration periods (defined as six weeks before and after the CNY holidays), and that migrant worker mobility patterns followed the patterns in the original mobility data during these migration periods. Because migration periods could span multiple holidays and inter-holiday periods, and the exact overlap varied by year due to the shifting dates of the CNY based on the lunar calendar, we estimated migrant worker flows based on the constructed traveler flows, which had already distributed the original mobility data according to the holiday schedule of each year. Specifically, for each year, we first estimated the total volume of migrant workers returning to their origin PLADs during the pre-CNY period, as well as the total volumes of recurring and new migrant workers moving to host PLADs during the post-CNY period. These estimates were calculated based on migrant worker population sizes for that year (see details in the Supplementary Text “Estimation of inter-PLAD rural-to-urban migrant worker population sizes”), the proportion of migrant workers remaining in host PLADs during CNY (32), the average duration of stay as migrant workers (33), and the employment rate of migrant workers (34) (see formulas in Eqs. S19– 22). Then, to estimate daily migrant worker volumes, the total migrant worker volumes were proportionally distributed based on the daily traveler volumes during the migration periods.

### Networked metapopulation SEIR model

The networked metapopulation SEIR model divided each calendar year into four periods based on migrant worker mobility patterns related to CNY: regular (no migration), pre-CNY (defined as six weeks before the CNY holidays; migrant workers return from host PLADs to origin PLADs), CNY holidays (one week; no migration), and post-CNY (six weeks after the CNY holidays; migrant workers move from origin PLADs to host PLADs) periods. PLADs ranked highest in both the proportion of migrant workers and per-capita GRP were determined as host PLADs in the model (seven host PLADs in total; Fig. S2). For each host PLAD, all other PLADs were ranked based on their proportions of the total migrant worker population, and the top-ranked PLADs collectively accounting for >75% of the total migrant worker population were determined as the origin PLADs for that host PLAD (18 origin PLADs in total; Fig. S2).

Jiangsu and Zhejiang served as both host and origin PLADs. The remaining eight PLADs were neither host nor origin PLADs. PLADs were further divided into subpopulations based on their host and origin roles. Each host PLAD was divided into a local subpopulation (including local residents and white-collar migrant workers with comparable population susceptibility) and migrant worker subpopulations (specifically rural-to-urban migrant workers) from corresponding origin PLADs (e.g., Beijing, Fig. 2b). Each origin PLAD was divided into a local subpopulation and returned migrant worker subpopulations (specifically rural-to-urban migrant workers) from corresponding host PLADs (e.g., Hebei, Fig. 2c). PLADs that served as both host and origin contained all three types of subpopulations (e.g., Jiangsu, Fig. 2a), and PLADs that were neither hosts nor origins contained only local subpopulations (e.g., Yunnan, Fig. 2d).

The model framework consisted of a basic SEIR model for each local, migrant worker, and returned migrant worker subpopulation in each PLAD. Subpopulations across PLADs were interconnected through a migrant worker network and a traveler network. Each basic SEIR model included transmission terms determined by population contact patterns and climate forcing (accounting for the effects of humidity and temperature on measles transmission), demographic processes (births and deaths), and routine childhood vaccination. Model formulation details are provided in the Supplementary Text “Networked metapopulation SEIR model”. Additionally, the nationwide SIA conducted in 2010 was modeled, and the details of the SIA modeling are provided in the Supplementary Text “Modeling nationwide SIA”.

The model was initialized with 2 million realizations generated using initialization methods for model state variables and Latin Hypercube sampling of model parameters (see initialization details in the Supplementary Text “Model initialization”). To calibrate the model, we first calculated the likelihood for each trajectory by comparing simulated incidence with observed incidence time series from 2005 to 2008 (a period with regular annual outbreaks), assuming a normal distribution centered at the observed incidence. We also filtered out trajectories whose proportions of incidence attributed to migrant worker subpopulations in host PLADs fell outside ranges reported in the literature (9, 11, 35–40). We then selected the 200 trajectories (i.e., the top 0.01%) with the highest likelihoods and resampled from these trajectories with replacement, using probability proportional to their likelihoods, to form a calibrated ensemble of 3,000 trajectories. The median estimates of calibrated model parameters are shown in Figs. S8 and S9. To simulate epidemic dynamics over the entire study period from 2005 to 2014 and to account for model stochasticity, each trajectory was reinitialized and rerun 10 times for this period. The resulting model ensemble served as the baseline for downstream analyses.

### Construction of counterfactual scenarios

Four counterfactual scenarios were constructed based on the networked metapopulation SEIR model. 1) No travelers: The traveler volumes in the travel network were set to 0; 2) No case importation: The volumes of exposed and infectious individuals in the migrant worker network were set to 0; 3) Matching population susceptibility: The population susceptibilities of migrant workers moving into host PLADs were set equal to those of the corresponding local subpopulations; 4) Pre-migration vaccination: We assumed that migrant workers were vaccinated with full coverage and 95% effectiveness prior to their departure to host PLADs. In the model, vaccination was implemented by transitioning susceptible individuals to the recovered/immunized compartment to achieve a population susceptibility of 5% among these migrant workers.

## Acknowledgement

This study was supported by the National Institute of Allergy and Infectious Diseases (AI145883).

## Data Availability Statement

The measles incidence data and the mobility data are subject to restriction. To access incidence data and/or to seek permission for its use, please contact the Data-center of China Public Health Science (https://www.phsciencedata.cn/Share/en/index.jsp). To access mobility data and/or to seek permission for its use, please contact Xiao-Ke Xu.

## Author Contribution

Peihua Wang: Conceptualization, Methodology, Formal Analysis, Writing–Original Draft; Xianwen Wang: Data curation, Validation, Writing–Review & Editing; Wenyi Zhang: Data curation, Formal Analysis, Validation, Writing–Review & Editing; Yong Wang: Data curation, Validation, Writing–Review & Editing; Sen Pei: Validation, Writing–Review & Editing; Xiao-Ke Xu: Data curation, Validation, Writing–Review & Editing; Wan Yang: Conceptualization, Methodology, Validation, Writing–Review & Editing, Funding acquisition, Supervision.

## Use of AI Statement

P.W. acknowledges the use of ChatGPT exclusively for grammar corrections during manuscript preparation.

