## Supplementary Information for "Rural-to-Urban Migrant Worker Mobility Shaped Measles Epidemics in China"

### Supplementary Figure

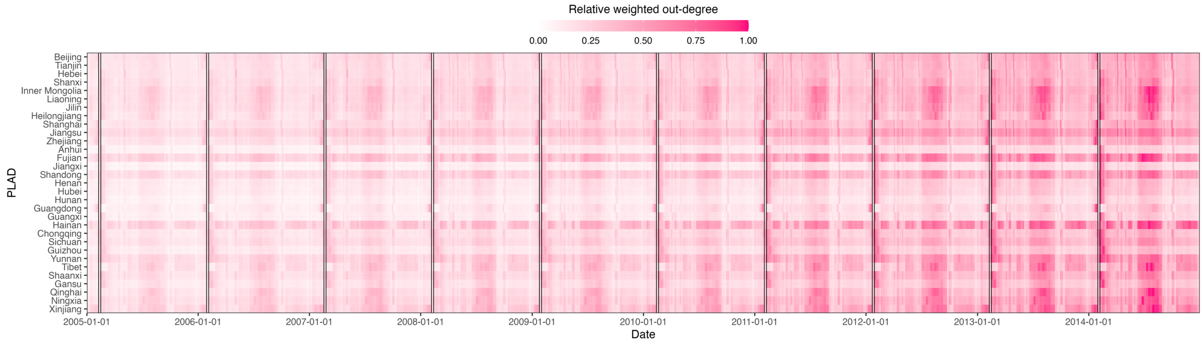


**Fig. S1** Relative weighted out-degree of the inter-PLAD traveler network from 2005 to 2014.

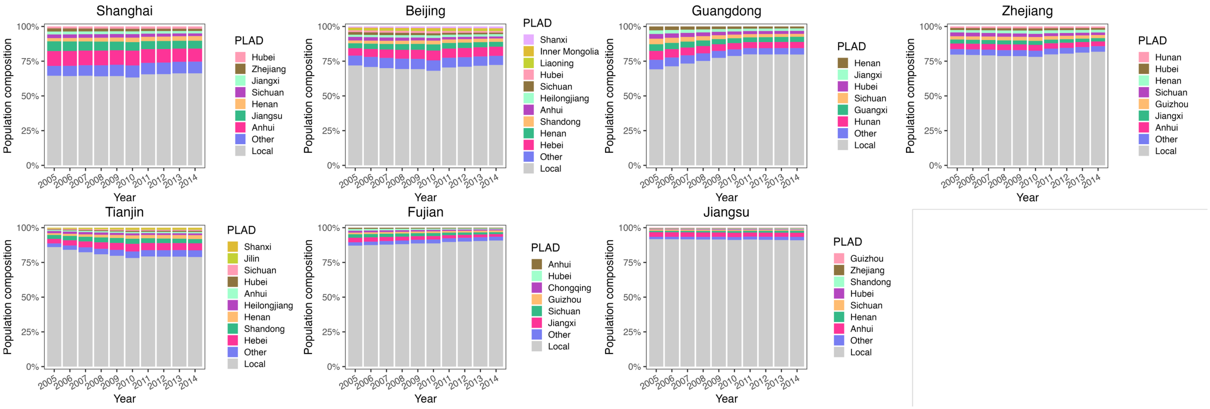


**Fig. S2** Population compositions of PLADs hosting the largest proportions of inter-PLAD rural-to-urban migrant workers (host PLADs). For each host PLAD, individual sources of migrant workers (origin PLADs) are ordered by their proportions from the smallest at the top to the largest at the bottom. They collectively account for >75% of the total migrant worker population.

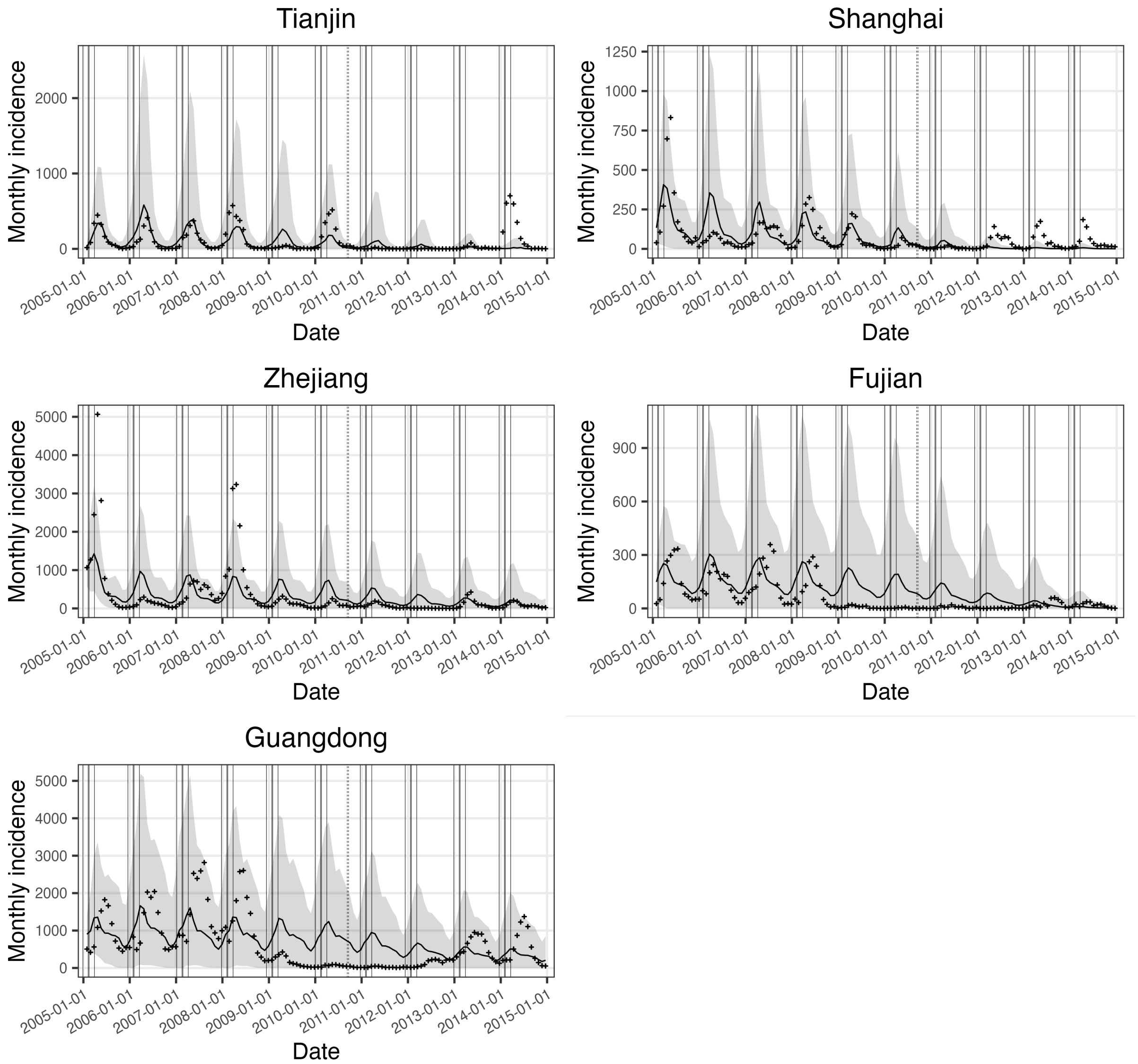


**Fig. S3** Simulated measles incidence from the calibrated model ensemble for host PLADs, compared with observed incidence (crosses).

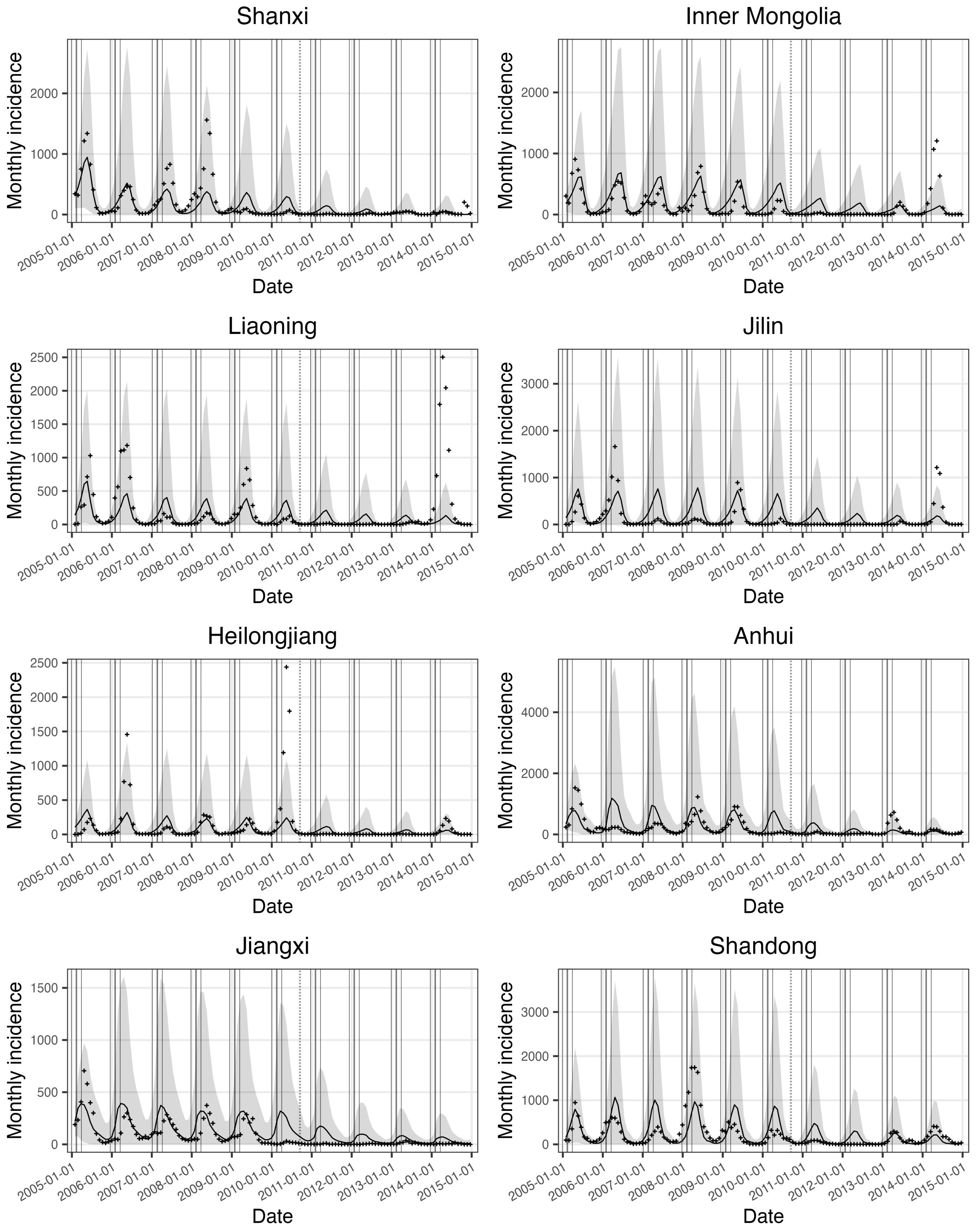


**Fig. S4** Simulated measles incidence from the calibrated model ensemble for origin PLADs, compared with observed incidence (crosses); part 1.

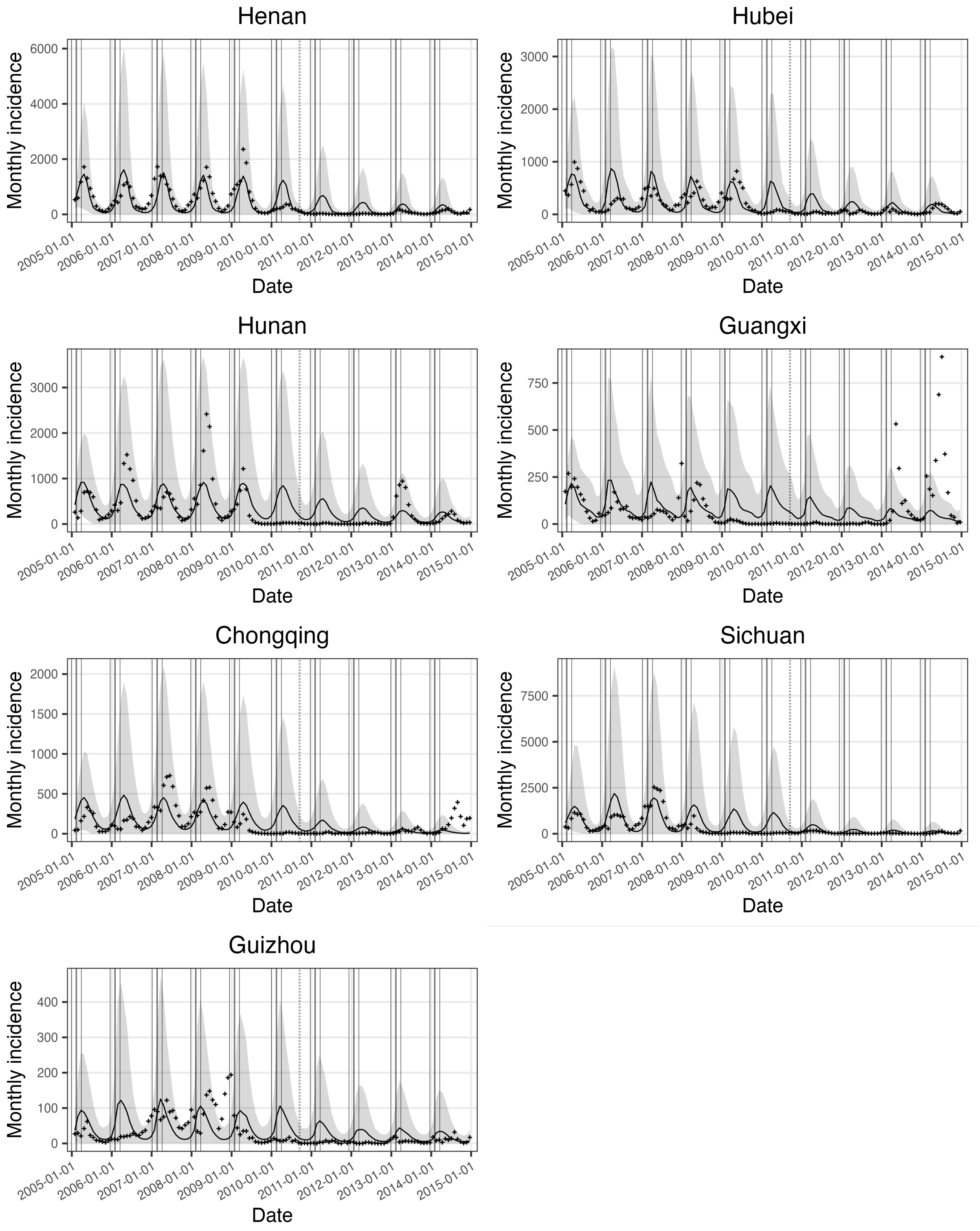


**Fig. S5** Simulated measles incidence from the calibrated model ensemble for origin PLADs, compared with observed incidence (crosses); part 2.

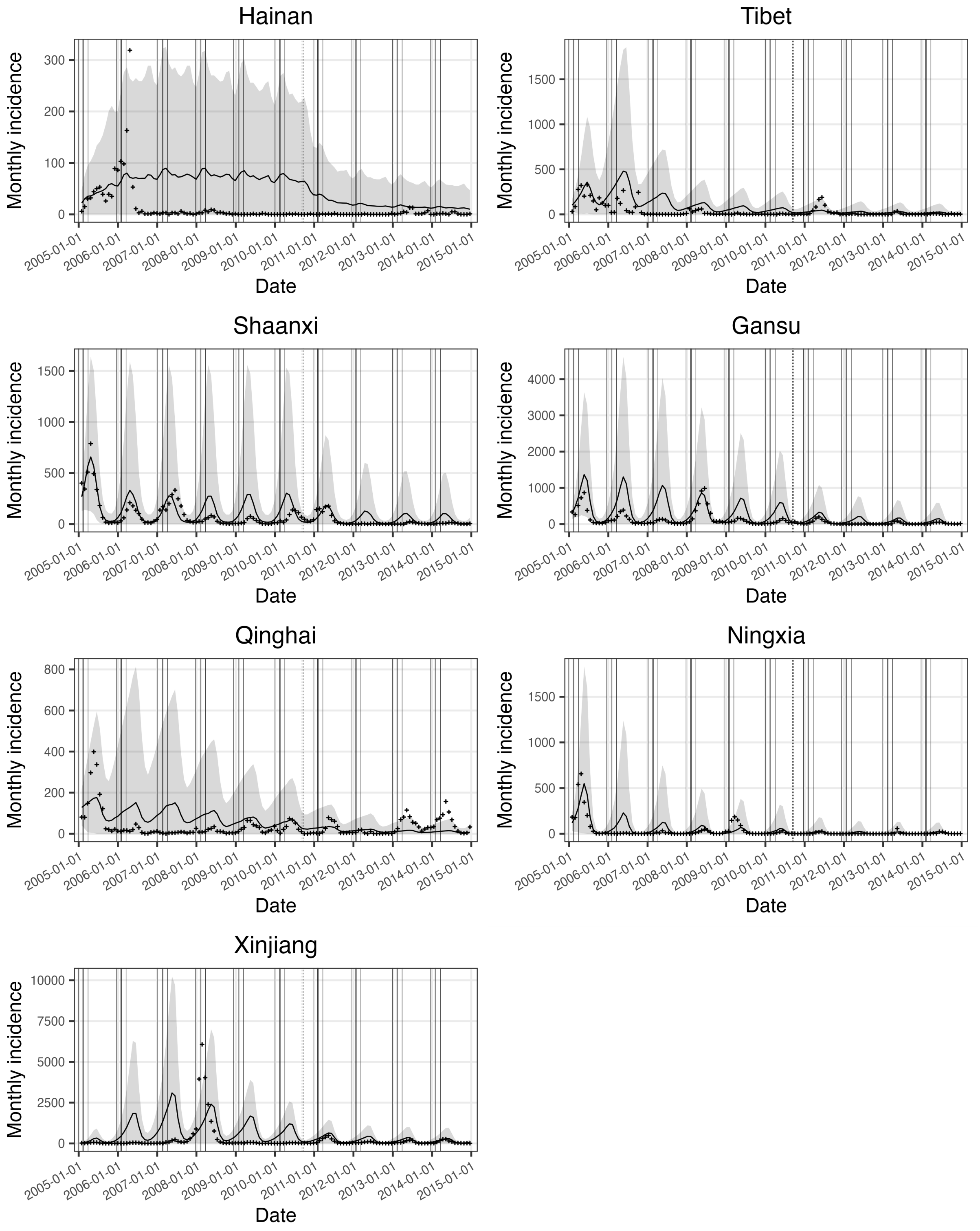


**Fig. S6** Simulated measles incidence from the calibrated model ensemble for PLADs neither hosts nor origins, compared with observed incidence (crosses).

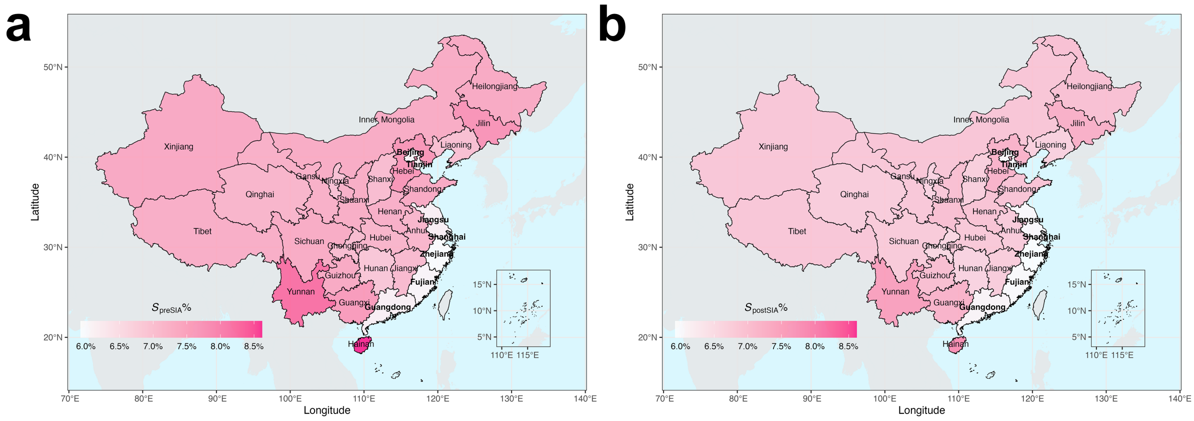


**Fig. S7** Median estimates of population susceptibilities (**a**) before and (**b**) after the nationwide SIA in 2010.

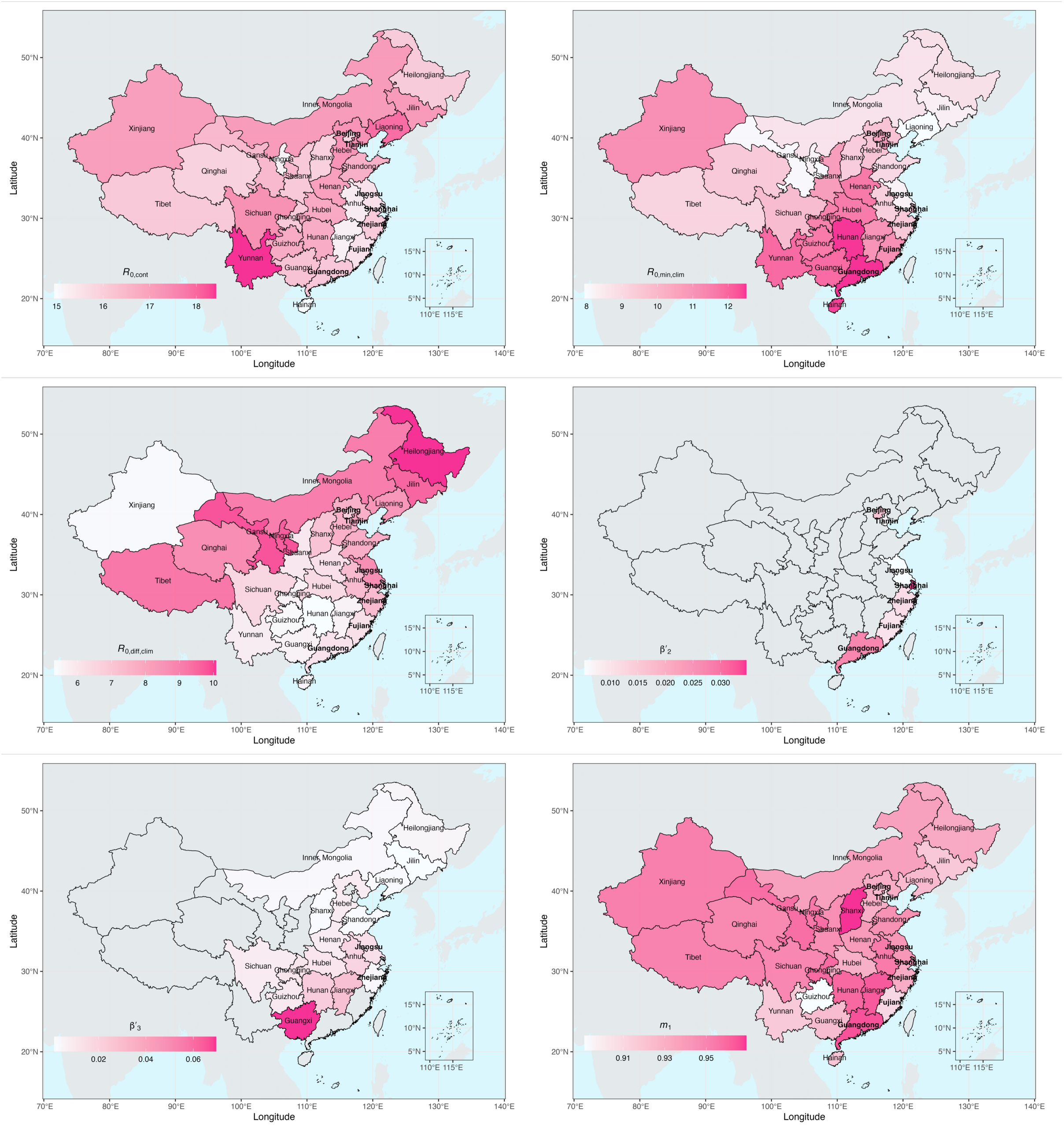


**Fig. S8** Median estimates of calibrated model parameters, part 1. Bolded fonts indicate host PLADs. Parameter definitions are provided in in the Supplementary Text "Networked metapopulation SEIR model".

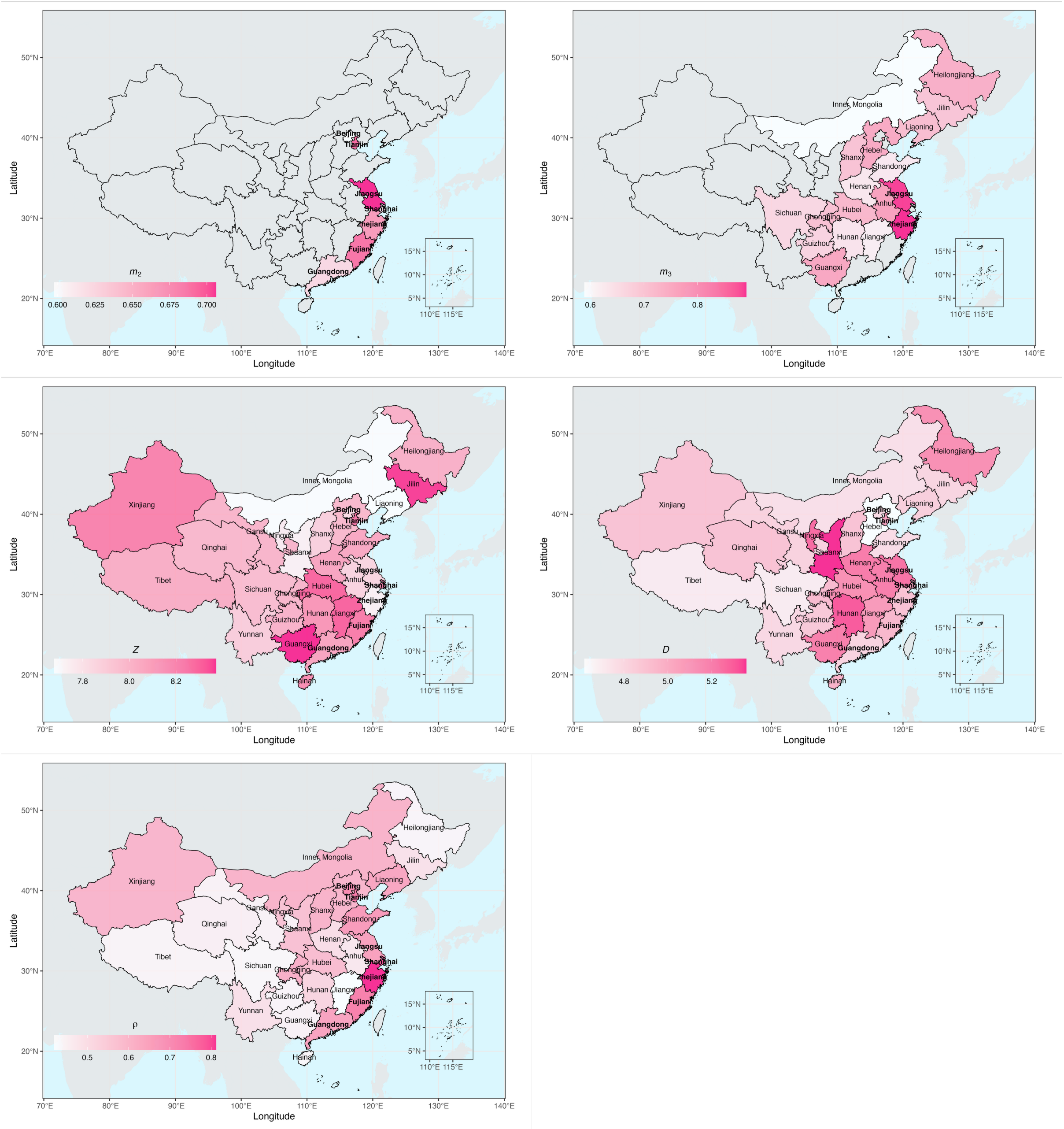


**Fig. S9** Median estimates of calibrated model parameters, part 2.

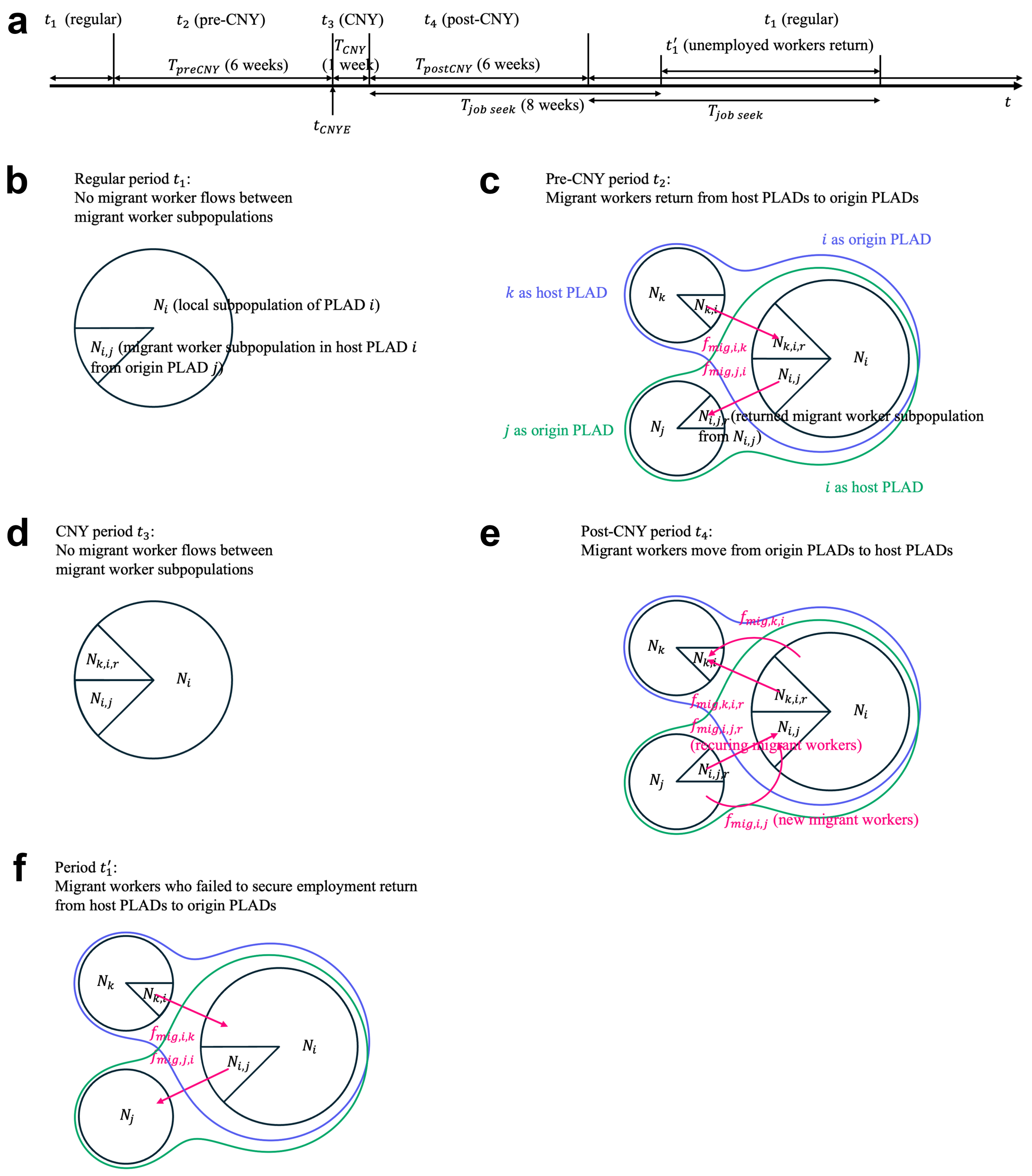


**Fig. S10** (**a**) Schematic of the defined time periods and their durations based on the migrant worker mobility patterns related to CNY in a typical year. $t_1$: regular period (no migration); $t_2$: pre-CNY migration period (duration $T_{preCNY}$: 6 weeks before the CNY's eve $t_{CNYE}$); $t_3$: CNY period (duration $T_{CNY}$: 1 week; no migration); $t_4$: post-CNY migration period (duration $T_{postCNY}$: 6 weeks); $t_1'$: period during which migrant workers who failed to secure employment return (duration of job seeking $T_{job\ seek}$: 8 weeks). Schematic of migrant worker flows between a focal PLAD $i$ serving as both a host and an origin PLAD (indicated by a larger pie), and connected PLADs $j$ and $k$ (indicated by smaller pies), during (**b**) $t_1$, (**c**) $t_2$, (**d**) $t_3$, (**e**) $t_4$, and (**f**) $t_1'$. For a focal host PLAD $i$, the migrant worker flows enclosed by green lines apply, and the returned migrant worker subpopulation $N_{k,i,r}$ should be ignored. For a focal origin PLAD $i$, the migrant worker flows enclosed by blue lines apply, and the migrant worker subpopulation $N_{i,j}$ should be ignored. A focal PLAD $i$ that is neither host nor origin only contains only a local subpopulation and has no migrant worker flows. Model details of migrant worker flows are provided in Eqs. 19–22.

**Supplementary Table**

**Table S1** Summary statistics of model fits for 2005–2008. RRMSE: Relative Root Mean Square Error; *r*: correlation coefficient; peak time difference: mean of absolute differences (in months) between observed and simulated peak timings.

| PLAD category | PLAD | RRMSE | *r* | Peak time difference |
|---|---|---|---|---|
| Host PLAD | Beijing | 0.397 | 0.935 | 1.00 |
| | Tianjin | 0.769 | 0.761 | 0.75 |
| | Shanghai | 1.101 | 0.570 | 1.25 |
| | Jiangsu | 0.728 | 0.725 | 0.25 |
| | Zhejiang | 1.308 | 0.641 | 1.00 |
| | Fujian | 0.753 | 0.414 | 2.75 |
| | Guangdong | 0.608 | 0.338 | 3.25 |
| Origin PLAD | Hebei | 1.137 | 0.874 | 0.25 |
| | Shanxi | 0.812 | 0.797 | 0.50 |
| | Neimenggu | 0.667 | 0.831 | 0.75 |
| | Liaoning | 1.314 | 0.623 | 0.50 |
| | Jilin | 1.980 | 0.508 | 0.50 |
| | Heilongjiang | 1.821 | 0.593 | 0.50 |
| | Anhui | 1.204 | 0.455 | 1.00 |
| | Jiangxi | 0.760 | 0.634 | 1.75 |
| | Shandong | 0.938 | 0.637 | 0.50 |

| | | | | |
|---|---|---|---|---|
| | Henan | 0.543 | 0.777 | 1.00 |
| | Hubei | 0.820 | 0.535 | 1.50 |
| | Hunan | 0.796 | 0.600 | 1.50 |
| | Guangxi | 0.901 | 0.350 | 1.50 |
| | Chongqing | 0.760 | 0.454 | 1.75 |
| | Sichuan | 0.799 | 0.647 | 0.25 |
| | Guizhou | 1.069 | -0.087 | 2.75 |
| PLAD neither host nor origin | Hainan | 2.633 | -0.132 | 1.50 |
| | Yunnan | 0.680 | 0.450 | 1.75 |
| | Tibet | 2.150 | 0.473 | 0.33 |
| | Shaanxi | 0.665 | 0.873 | 0.50 |
| | Gansu | 1.916 | 0.661 | 0.25 |
| | Qinghai | 2.112 | 0.596 | 0.33 |
| | Ningxia | 1.801 | 0.771 | 0.67 |
| | Xinjiang | 3.000 | 0.249 | 1.50 |

**Table S2** Relative differences in cumulative incidence between counterfactual scenarios and the baseline in origin PLADs.

| Origin PLAD | Counterfactual scenario | | | |
|---|---|---|---|---|
| | No travelers | No case importation | Matching population susceptibility | Pre-migration vaccination |
| Hebei | 2.0% (-21.5%, 28.8%) | -20.4% (-39.5%, 3.2%) | -15.0% (-36.2%, 9.3%) | -34.9% (-50.2%, -16.8%) |
| Shanxi | 4.1% (-27.5%, 46.8%) | -2.3% (-32.5%, 36.9%) | -1.6% (-31.7%, 34.0%) | -9.3% (-36.1%, 24.3%) |

| | | | | |
|---|---|---|---|---|
| Inner Mongolia | 2.1% (-28.8%, 38.4%) | -2.1% (-29.6%, 35.5%) | 0.4% (-29.9%, 37.8%) | -8.1% (-34.2%, 24.5%) |
| Liaoning | -1.3% (-30.4%, 37.3%) | -1.4% (-32.7%, 36.7%) | -0.7% (-30.2%, 38.6%) | -6.7% (-35.8%, 28.5%) |
| Jilin | -4.4% (-31.3%, 31.8%) | -0.7% (-29.6%, 36.5%) | -0.3% (-28.4%, 36.5%) | -3.0% (-31.7%, 31.0%) |
| Heilongjiang | 1.2% (-28.4%, 39.8%) | -10.4% (-38.5%, 28.0%) | -7.9% (-36.1%, 29.0%) | -21.3% (-44.4%, 9.6%) |
| Anhui | 1.3% (-29.1%, 39.7%) | -34.5% (-65.1%, -0.7%) | -7.0% (-43.2%, 34.0%) | -69.9% (-79.6%, -57.7%) |
| Jiangxi | -1.5% (-26.2%, 28.7%) | -50.7% (-65.5%, -32.1%) | -21.1% (-43.3%, 5.8%) | -71.9% (-78.6%, -64.0%) |
| Shandong | 1.1% (-23.1%, 29.6%) | -19.0% (-40.6%, 7.2%) | -14.9% (-36.0%, 11.5%) | -29.0% (-47.8%, -6.7%) |
| Henan | -0.9% (-22.1%, 24.3%) | -36.3% (-53.1%, -16.9%) | -21.4% (-40.7%, 0.2%) | -59.1% (-69.0%, -48.1%) |
| Hubei | -2.7% (-27.0%, 26.5%) | -41.0% (-59.7%, -18.1%) | -21.8% (-44.4%, 5.8%) | -65.4% (-75.1%, -54.4%) |
| Hunan | -2.2% (-24.2%, 24.8%) | -21.7% (-42.6%, 6.4%) | -2.5% (-26.8%, 28.0%) | -61.8% (-71.2%, -49.8%) |
| Guangxi | -3.1% (-26.5%, 24.6%) | -47.1% (-62.6%, -28.0%) | -23.0% (-43.0%, 3.3%) | -68.7% (-76.7%, -58.7%) |
| Chongqing | 5.9% (-24.3%, 44.7%) | -2.3% (-30.0%, 34.0%) | 1.7% (-28.5%, 40.3%) | -13.2% (-39.0%, 20.1%) |
| Sichuan | 3.7% (-23.7%, 36.0%) | -35.4% (-54.6%, -13.8%) | -16.3% (-40.9%, 12.7%) | -62.6% (-73.2%, -50.5%) |
| Guizhou | 2.4% (-21.6%, 30.6%) | -57.2% (-68.4%, -45.4%) | -40.7% (-55.7%, -24.7%) | -65.6% (-73.5%, -56.3%) |

**Table S3** Relative differences in cumulative incidence between counterfactual scenarios and the baseline in PLADs that were neither host nor origin PLADs.

| Other PLAD | Counterfactual scenario | | | |
|---|---|---|---|---|
| | No travelers | No case importation | Matching population susceptibility | Pre-migration vaccination |
| Hainan | 0.3% (-24.9%, 32.3%) | 1.5% (-24.1%, 31.2%) | -0.1% (-24.4%, 28.1%) | 2.3% (-22.6%, 31.9%) |
| Yunnan | 4.1% (-4.6%, 13.7%) | 0.2% (-8.5%, 9.3%) | 0.0% (-8.0%, 8.8%) | 0.3% (-8.2%, 9.0%) |

| | | | | |
|---|---|---|---|---|
| Tibet | -3.9% (-22.9%, 18.2%) | 0.7% (-20.1%, 23.3%) | 0.5% (-18.4%, 23.4%) | 1.9% (-18.3%, 25.5%) |
| Shaanxi | -12.7% (-31.5%, 11.0%) | 0.4% (-23.2%, 27.4%) | 2.2% (-19.6%, 27.6%) | 3.5% (-18.6%, 31.4%) |
| Gansu | 4.1% (-14.2%, 26.0%) | -0.1% (-17.3%, 20.1%) | -0.1% (-17.8%, 19.3%) | -3.8% (-20.8%, 15.2%) |
| Qinghai | 2.0% (-28.0%, 39.1%) | 0.7% (-27.4%, 36.7%) | 1.5% (-26.4%, 37.1%) | -4.6% (-31.4%, 28.8%) |
| Ningxia | -4.3% (-30.8%, 28.2%) | -0.1% (-27.2%, 35.5%) | -0.1% (-27.1%, 32.7%) | -0.2% (-27.7%, 35.3%) |
| Xinjiang | 2.9% (-13.0%, 21.4%) | 0.4% (-15.3%, 17.3%) | 0.8% (-13.8%, 18.4%) | 1.1% (-13.8%, 19.0%) |

**Table S4** Values of constant model parameters.

| Parameter | Value |
|---|---|
| Proportion of migrant workers who remain in host PLADs during CNY, $\theta$ | 0.5 (1) |
| Employment rate of migrant workers, $\eta$ | 0.7 (2) |
| Average duration of stay as migrant workers, $L$ (year) | 3 (3) |
| Duration of return migration to origin PLADs before CNY, $T_{preCNY}$ (week) | 6 |
| Duration of CNY holiday, $T_{CNY}$ (week) | 1 |
| Duration of migration to host PLADs after CNY, $T_{postCNY}$ (week) | 6 |
| Duration of job-seeking period of migrant workers in host PLADs, $T_{job\ seek}$ (week) | 8 |

**Table S5** Initial ranges of model state variables and parameters.

| State variable and parameter | | Range |
|---|---|---|
| | Susceptible population ($S$) | Population model (see Supplementary Text "Model initialization") |

| | | |
|---|---|---|
| State variable | Exposed population ($E$) | Initial observed incidence adjusted by reporting rate, distributed among subpopulations assuming equal prevalence, and sampled from a negative binomial distribution |
| | Infectious population ($I$) | Initial observed incidence adjusted by reporting rate, distributed among subpopulations assuming equal prevalence, and sampled from a negative binomial distribution |
| Parameter | Basic reproductive number based on contact ($R_{0,cont}$) | [8, 20] (4) |
| | Minimum daily basic reproductive number in an absolute humidity and temperature-forced model ($R_{0,min,clim}$) | [6, 15] (4) |
| | Difference between maximum and minimum daily basic reproductive numbers in an absolute humidity and temperature-forced model ($R_{0,diff,clim}$) | [4, 18] (4) |
| | Transmission rate within subpopulation, relative to $\beta_1$ ($\beta_1'$) | 1 |
| | Transmission rate between subpopulations originally from different PLADs, relative to $\beta_1$ ($\beta_2'$) | $[0, \frac{0.4}{a}]$ (the upper bound (sum of the element (1,2) in Eq. 18) was set as 0.4, estimated by the largest proportion of migrant workers among PLADs, and by |

| | | |
|---|---|---|
| | | assuming homogeneous mixing among local and migrant worker subpopulations) |
| | Transmission rate between subpopulations originally from a same PLAD, relative to $\beta_1$ ($\beta_3'$) | $[0, \frac{0.2}{b}]$ (the upper bound (sum of the element (1,3) in Eq. 18) was set as 0.2, estimated by the largest proportion of returning migrant workers among PLADs, and by assuming the local subpopulation had the same contact rates within itself and with the returning migrant worker subpopulations) |
| | Mixing exponent within subpopulation ($m_1$) | [0.85, 1] |
| | Mixing exponent between subpopulations originally from different PLADs ($m_2$) | [0.4, 1] |
| | Mixing exponent between subpopulations originally from a same PLAD ($m_3$) | [0.4, 1] |
| | Latent period ($Z$) | [7, 9] (4) |
| | Infectious period ($D$) | [4, 6] (4) |
| | Reporting rate ($\rho$) | Time series susceptible–infected–recovered model (4-6) |

### Supplementary Text

**Estimation of inter-PLAD rural-to-urban migrant worker population sizes.** Population sizes of inter-PLAD migrant workers (represented as a contingency matrix where rows indicated host PLADs, columns indicated origin PLADs, and diagonal elements were 0) were reported in the national population sample surveys in 2005 (7) and 2015 (8), and in the national census in 2010 (9). Annual total inter-PLAD rural-to-urban migrant worker population sizes from 2005 to 2014 were reported by the National Bureau of Statistics of China (10). To estimate inter-PLAD rural-to-urban migrant worker population sizes from 2005 to 2014, we assumed that: 1) the population size of inter-PLAD rural-to-urban migrant workers for each host-origin PLAD pair scaled proportionally to the annual total inter-PLAD migrant worker population size; and 2) the proportion of the rural-to-urban migrant worker population size for each host-origin PLAD pair relative to the total rural-to-urban migrant worker population size changed linearly between survey years. Specifically, we first normalized the contingency matrices of inter-PLAD migrant worker populations for the survey years 2005, 2010, and 2015. We then estimated normalized contingency matrices for the intermediate years (2006–2009 and 2011–2014) using linear interpolation. Lastly, inter-PLAD rural-to-urban migrant worker population sizes from 2005 to 2014 were computed by multiplying these contingency matrices by the corresponding annual total inter-PLAD rural-to-urban migrant worker population sizes.

**Networked metapopulation SEIR model.** The model divided each calendar year into four periods based on the migrant worker mobility patterns related to CNY: regular ($t_1$), pre-CNY ($t_2$), CNY ($t_3$), and post-CNY ($t_4$) periods (see schematic in Fig. S10a):

$$t_1 \in [t_{Jan\,01}, t_{CNYE} - T_{preCNY}] \cup (t_{CNYE} + T_{CNY} + T_{postCNY}, t_{Dec\,31}] \quad (1)$$

$$t_1' \subseteq t_1, t_1' \in (t_{CNYE} + T_{CNY} + T_{job\,seek}, t_{CNYE} + T_{CNY} + T_{postCNY} + T_{job\,seek}]$$

$$t_2 \in \left(t_{CNYE} - T_{preCNY}, t_{CNYE}\right]$$

$$t_3 \in (t_{CNYE}, t_{CNYE} + T_{CNY}]$$

$$t_4 \in \left(t_{CNYE} + T_{CNY}, t_{CNYE} + T_{CNY} + T_{postCNY}\right]$$

Here, $t_1'$ is a sub-period within $t_1$, during which migrant workers who did not secure employment in host PLADs returned to their origin PLADs. $t_{CNYE}$ is the date of CNY's Eve, and $T$ is the duration of each time period (see parameter values in Table S3).

A PLAD could be divided into up to three types of subpopulations. A PLAD $i$ serving as both a host and an origin (e.g., Jiangsu) contained all three types of subpopulations:

1) a local subpopulation ($N_i$ in Figs. S10b–f; its population dynamics corresponding to, e.g., "Jiangsu" in Fig. 3a);

2) migrant worker subpopulations from corresponding origin PLADs ($N_{i,j}$, in Figs. S10b–f, where $j \in A$, and $A$ is the set of corresponding origin PLADs; e.g., "Jiangsu_Anhui" in Fig. 3a, $A$ = {Anhui, Henan, Sichuan, Hubei, Shandong, Zhejiang, Guizhou});

3) returned migrant worker subpopulations from corresponding host PLADs ($N_{k,i,r}$ in Figs. S10b–f, where $k \in B$, $B$ is the set of corresponding host PLADs, and $r$ indicates returned subpopulation; e.g., "Shanghai_Jiangsu_ r" in Fig. 3a, $B$ = {Shanghai}).

Accordingly, a host PLAD contained a local subpopulation and migrant worker subpopulations (e.g., Beijing, Fig. 3b), an origin PLAD contained a local subpopulation and returned migrant worker subpopulations (e.g., Hebei, Fig. 3c), and a PLAD neither host nor origin contained only the local subpopulation (e.g., Yunnan, Fig. 3d).

Measles transmission dynamics within each subpopulation in PLAD $i$ were simulated using a metapopulation SEIR model integrated with a migrant worker network and a traveler network, with a daily time step.

For the local subpopulation $N_i$, the dynamics is governed by the following differential equations:

$$\begin{aligned}\frac{dS_i}{dt} = & -S_i\left(\beta_{1,i}(t)\frac{I_i^{m_{1,i}}}{N_i} + \sum_{a\in A}\beta_{2,i}(t)\frac{I_{i,a}^{m_{2,i}}}{N_{i,a}} + \sum_{b\in B}\beta_{3,i}(t)\frac{I_{b,i,r}^{m_{3,i}}}{N_{b,i,r}}\right) \\ & + \sum_{c\in C}\left(q_{i,c}(t)\frac{S_c}{N_c} + \left(1-q_{i,c}(t)\right)\frac{S_i}{N_i}\right)\left(f_{trav,i,c}(t) - f_{trav,c,i}(t)\right) \\ & - 1_{\{t\in t_4\}}\sum_{b\in B}\frac{S_i}{N_i}f_{mig,b,i}(t) + 1_{\{t\in t_1'\}}\sum_{b\in B}\frac{S_{b,i}}{N_{b,i}}f_{mig,i,b}(t) + f_{migo,i}(t)\frac{S_i}{N_i} \\ & + \lambda_i(t)N_i\left(1-\xi_i(t)\right) - \mu_i(t)S_i\end{aligned} \quad (2)$$

Structure of susceptible population dynamics: terms sequentially represent transmission from within the local subpopulation, from migrant worker subpopulations, from returned migrant worker subpopulations, traveler mobility, migrant worker mobility ($t \in t_4 \cup t_1'$), white-collar migrant worker mobility, unimmunized births, and deaths.

$$\frac{dE_i}{dt} = S_i\left(\beta_{1,i}(t)\frac{I_i^{m_{1,i}}}{N_i} + \sum_{a\in A}\beta_{2,i}(t)\frac{I_{i,a}^{m_{2,i}}}{N_{i,a}} + \sum_{b\in B}\beta_{3,i}(t)\frac{I_{b,i,r}^{m_{3,i}}}{N_{b,i,r}}\right) - \frac{E_i}{Z} + \sum_{c\in C}\left(q_{i,c}(t)\frac{E_c}{N_c} + \left(1 - q_{i,c}(t)\right)\frac{E_i}{N_i}\right)\left(f_{trav,i,c}(t) - f_{trav,c,i}(t)\right) - 1_{\{t\in t_4\}}\sum_{b\in B}\frac{E_i}{N_i}f_{mig,b,i}(t) + 1_{\{t\in t_1'\}}\sum_{b\in B}\frac{E_{b,i}}{N_{b,i}}f_{mig,i,b}(t) + f_{migo,i}(t)\frac{E_i}{N_i} - \mu_i(t)E_i \quad (3)$$

Structure of exposed population dynamics: terms sequentially represent transmission from within the local subpopulation, from migrant worker subpopulations, from returned migrant worker subpopulations, transition from exposed to infectious, traveler mobility, migrant worker mobility ($t \in t_4 \cup t_1'$), white-collar migrant worker mobility, and deaths.

$$\frac{dI_i}{dt} = \frac{E_i}{Z} - \frac{I_i}{D} + \sum_{c\in C}\left(q_{i,c}(t)\frac{I_c}{N_c} + \left(1 - q_{i,c}(t)\right)\frac{I_i}{N_i}\right)\left(f_{trav,i,c}(t) - f_{trav,c,i}(t)\right) - 1_{\{t\in t_4\}}\sum_{b\in B}\frac{I_i}{N_i}f_{mig,b,i}(t) + 1_{\{t\in t_1'\}}\sum_{b\in B}\frac{I_{b,i}}{N_{b,i}}f_{mig,i,b}(t) + f_{migo,i}(t)\frac{I_i}{N_i} - \mu_i(t)I_i \quad (4)$$

Structure of infectious population dynamics: terms sequentially represent transition from exposed to infectious, transition from infectious to recovered, traveler mobility, migrant worker mobility ($t \in t_4 \cup t_1'$), white-collar migrant worker mobility, and deaths.

$$\frac{dN_i}{dt} = \sum_{c\in C}\left(f_{trav,i,c}(t) - f_{trav,c,i}(t)\right) - 1_{\{t\in t_4\}}\sum_{b\in B}f_{mig,b,i}(t) + 1_{\{t\in t_1'\}}\sum_{b\in B}f_{mig,i,b}(t) + f_{migo,i}(t) + (\lambda_i(t) - \mu_i(t))N_i \quad (5)$$

For a migrant worker subpopulation $N_{i,j}$ in PLAD $i$:

$$\frac{dS_{i,j}}{dt} = -S_{i,j}(\beta_{1,i}(t)\frac{I_{i,j}^{m_{1,i}}}{N_{i,j}} + \beta_{2,i}(t)\frac{I_i^{m_{2,i}}}{N_i} + \sum_{a\in A, a\neq j}\beta_{2,i}(t)\frac{I_{i,a}^{m_{2,i}}}{N_{i,a}} + \sum_{b\in B}\beta_{2,i}(t)\frac{I_{b,i,r}^{m_{2,i}}}{N_{b,i,r}}) \tag{6}$$
$$- 1_{\{t\in t_2\}}\frac{S_{i,j}}{N_{i,j}}f_{mig,j,i}(t) + 1_{\{t\in t_4\}}\left(\frac{S_{i,j,r}}{N_{i,j,r}}f_{mig,i,j,r}(t) + \frac{S_j}{N_j}f_{mig,i,j}(t)\right)$$
$$- 1_{\{t\in t_1'\}}\frac{S_{i,j}}{N_{i,j}}f_{mig,j,i}(t) + \lambda_i(t)N_{i,j}(1-\xi_i(t)) - \mu_i(t)S_{i,j}$$

Structure of susceptible population dynamics: terms sequentially represent transmission from within the migrant worker subpopulation, from local subpopulation, from other migrant worker subpopulations, from returned migrant worker subpopulations, migrant worker mobility ($t \in t_2 \cup t_4 \cup t_1'$), unimmunized births, and deaths.

$$\frac{dE_{i,j}}{dt} = S_{i,j}\left(\beta_{1,i}(t)\frac{I_{i,j}^{m_{1,i}}}{N_{i,j}} + \beta_{2,i}(t)\frac{I_i^{m_{2,i}}}{N_i} + \sum_{a\in A, a\neq j}\beta_{2,i}(t)\frac{I_{i,a}^{m_{2,i}}}{N_{i,a}} + \sum_{b\in B}\beta_{2,i}(t)\frac{I_{b,i,r}^{m_{2,i}}}{N_{b,i,r}}\right) \tag{7}$$
$$- \frac{E_{i,j}}{Z} - 1_{\{t\in t_2\}}\frac{E_{i,j}}{N_{i,j}}f_{mig,j,i}(t)$$
$$+ 1_{\{t\in t_4\}}\left(\frac{E_{i,j,r}}{N_{i,j,r}}f_{mig,i,j,r}(t) + \frac{E_j}{N_j}f_{mig,i,j}(t)\right) - 1_{\{t\in t_1'\}}\frac{E_{i,j}}{N_{i,j}}f_{mig,j,i}(t)$$
$$- \mu_i(t)E_{i,j}$$

Structure of exposed population dynamics: terms sequentially represent transmission from within the migrant worker subpopulation, from local subpopulation, from other migrant worker subpopulations, from returned migrant worker subpopulations, transition from exposed to infectious, migrant worker mobility ($t \in t_2 \cup t_4 \cup t_1'$), and deaths.

$$\frac{dI_{i,j}}{dt} = \frac{E_{i,j}}{Z} - \frac{I_{i,j}}{D} - 1_{\{t\in t_2\}}\frac{I_{i,j}}{N_{i,j}}f_{mig,j,i}(t) \tag{8}$$
$$+ 1_{\{t\in t_4\}}\left(\frac{I_{i,j,r}}{N_{i,j,r}}f_{mig,i,j,r}(t) + \frac{I_j}{N_j}f_{mig,i,j}(t)\right) - 1_{\{t\in t_1'\}}\frac{I_{i,j}}{N_{i,j}}f_{mig,j,i}(t)$$
$$- \mu_i(t)I_{i,j}$$

Structure of infectious population dynamics: terms sequentially represent transition from exposed to infectious, transition from infectious to recovered, migrant worker mobility ($t \in t_2 \cup t_4 \cup t_1'$), and deaths.

$$\frac{dN_{i,j}}{dt} = -1_{\{t\in t_2\}} f_{mig,j,i}(t) + 1_{\{t\in t_4\}}\left(f_{mig,i,j,r}(t) + f_{mig,i,j}(t)\right) - 1_{\{t\in t_1'\}} f_{mig,j,i}(t) + (\lambda_i(t) - \mu_i(t))N_{i,j} \tag{9}$$

For a returned migrant worker subpopulation $N_{k,i,r}$ in PLAD $i$:

$$\frac{dS_{k,i,r}}{dt} = -S_{k,i,r}\left(\beta_{1,i}(t)\frac{I_{k,i,r}^{m_{1,i}}}{N_{k,i,r}} + \beta_{3,i}(t)\frac{I_i^{m_{3,i}}}{N_i} + \sum_{a\in A}\beta_{2,i}(t)\frac{I_{i,a}^{m_{2,i}}}{N_{i,a}} + \sum_{b\in B, b\neq k}\beta_{3,i}(t)\frac{I_{b,i,r}^{m_{3,i}}}{N_{b,i,r}}\right) + 1_{\{t\in t_2\}}\frac{S_{k,i}}{N_{k,i}} f_{mig,i,k}(t) - 1_{\{t\in t_4\}}\frac{S_{k,i,r}}{N_{k,i,r}} f_{mig,k,i,r}(t) + \lambda_i(t)N_{k,i,r}\left(1-\xi_i(t)\right) - \mu_i(t)S_{k,i,r} \tag{10}$$

Structure of susceptible population dynamics: terms sequentially represent transmission from within the returned migrant worker subpopulation, from local subpopulation, from migrant worker subpopulations, from other returned migrant worker subpopulations, migrant worker mobility ($t \in t_2 \cup t_4$), unimmunized births, and deaths.

$$\frac{dE_{k,i,r}}{dt} = S_{k,i,r}\left(\beta_{1,i}(t)\frac{I_{k,i,r}^{m_{1,i}}}{N_{k,i,r}} + \beta_{3,i}(t)\frac{I_i^{m_{3,i}}}{N_i} + \sum_{a\in A}\beta_{2,i}(t)\frac{I_{i,a}^{m_{2,i}}}{N_{i,a}} + \sum_{b\in B, b\neq k}\beta_{3,i}(t)\frac{I_{b,i,r}^{m_{3,i}}}{N_{b,i,r}}\right) - \frac{E_{k,i,r}}{Z} + 1_{\{t\in t_2\}}\frac{E_{k,i}}{N_{k,i}} f_{mig,i,k}(t) - 1_{\{t\in t_4\}}\frac{E_{k,i,r}}{N_{k,i,r}} f_{mig,k,i,r}(t) - \mu_i(t)E_{k,i,r} \tag{11}$$

Structure of exposed population dynamics: terms sequentially represent transmission from within the returned migrant worker subpopulation, from local subpopulation, from migrant worker subpopulations, from other returned migrant worker subpopulations, transition from exposed to infectious, migrant worker mobility ($t \in t_2 \cup t_4$), and deaths.

$$\frac{dI_{k,i,r}}{dt} = \frac{E_{k,i,r}}{Z} - \frac{I_{k,i,r}}{D} + 1_{\{t\in t_2\}}\frac{I_{k,i}}{N_{k,i}} f_{mig,i,k}(t) - 1_{\{t\in t_4\}}\frac{I_{k,i,r}}{N_{k,i,r}} f_{mig,k,i,r}(t) - \mu_i(t) I_{k,i,r} \tag{12}$$

Structure of infectious population dynamics: terms sequentially represent transition from exposed to infectious, transition from infectious to recovered, migrant worker mobility ($t \in t_2 \cup t_4$), and deaths.

$$\frac{dN_{k,i,r}}{dt} = 1_{\{t\in t_2\}} f_{mig,i,k}(t) - 1_{\{t\in t_4\}} f_{mig,k,i,r}(t) + \big(\lambda_i(t) - \mu_i(t)\big) N_{k,i,r} \tag{13}$$

At $t = t_{CNYE} + T_{CNY} + T_{postCNY}$ (the end of post-CNY period $t_3$), returned migrant workers who remained in their origin PLADs and were no longer seeking employment in host PLADs were merged into the local subpopulation:

$$S_i = S_i + \sum_{b\in B} S_{b,i,r}\,, E_i = E_i + \sum_{b\in B} E_{b,i,r}\,, I_i = I + \sum_{b\in B} I_{b,i,r}\,, N_i = N_i + \sum_{b\in B} N_{b,i,r} \tag{14}$$

$$S_{b,i,r} = E_{b,i,r} = I_{b,i,r} = N_{b,i,r} = 0, b \in B$$

In Eqs. S2–13, we defined $\frac{x}{N_{b,i,r}} := \begin{cases} \frac{x}{N_{b,i,r}}, N_{b,i,r} \neq 0 \\ 0, N_{b,i,r} = 0 \end{cases}$, where $x$ is an arbitrary real number and $b \in B$, to set the transmission dynamics to 0 for the returned migrant worker subpopulation that has been merged into the local subpopulation. The parts of the equations shown in black represent the basic SEIR model, including transmission terms, demographic processes (birth and death), and routine childhood vaccination. $S$, $E$, $I$, and $N$ are the susceptible, exposed, infectious, and total populations, respectively, with $R = N - S - E - I$ representing those recovered and/or immunized. $\beta_1$ is the transmission rate within each subpopulation. $\beta_2$ is the transmission rate between subpopulations originating from different PLADs. Specifically, in PLAD $i$, $\beta_2$ applies to interactions between a migrant worker subpopulation $N_{i,j}$ (from $j$) and the local subpopulation $N_i$ (from $i$), between $N_{i,j}$ (from $j$) and a returned migrant worker subpopulation $N_{k,i,r}$ (from $i$), and between $N_{i,j}$ (from $j$) and another migrant worker subpopulation $N_{i,l}$ (from $l$). $\beta_3$ is the transmission rate between subpopulations originating from the same PLAD, specifically, between the local subpopulation $N_i$ (from $i$) and a returned migrant worker subpopulation $N_{k,i,r}$ (from $k$), and between two returned migrant worker subpopulations $N_{k,i,r}$ and $N_{o,i,r}$ (both from $i$). $m_1$, $m_2$, and $m_3$ indicate the degrees of inhomogeneous mixing, and their subscripts correspond to those defined by $\beta_1$, $\beta_2$, and $\beta_3$, respectively. $Z$ and $D$ are the latent and infectious periods, respectively. $\lambda$ and $\mu$ are the birth and death rates, respectively. $\xi$ is the immunization rate of routine childhood vaccination. Transition rates between model state variables (i.e., $S$, $E$, $I$, and $R$) were drawn from Poisson distributions to simulate transmission stochasticity.

In the basic SEIR model, the transmission rates in PLAD $i$ ($\beta_{x,i}(t)$) were calculated using the next generation matrix method (11):

$$\beta_{x,i}(t) = \beta'_{x,i} \frac{R_{0,cont,i}}{\rho(\boldsymbol{K}_i)} \frac{R_{0,clim,i}(t)}{\overline{R_{0,clim,i}}}, x \in \{1,2,3\} \quad (15)$$

$$\boldsymbol{K}_i = \boldsymbol{N}_i \boldsymbol{\beta}'_i \boldsymbol{N}_i^{-1} D \quad (16)$$

$$\boldsymbol{N}_i = \begin{bmatrix} N_i & \boldsymbol{0}_{|A|}^{\top} & \boldsymbol{0}_{|B|}^{\top} \\ \boldsymbol{0}_{|A|} & diag\{N_{i,a}\}_{a\in A} & \boldsymbol{0}_{|A|,|B|} \\ \boldsymbol{0}_{|B|} & \boldsymbol{0}_{|B|,|A|} & diag\{N_{b,i,r}\}_{b\in B} \end{bmatrix} \quad (17)$$

$$\boldsymbol{\beta}'_i = \begin{bmatrix} \beta'_{1,i} & \beta'_{2,i}\boldsymbol{1}_{|A|}^{\top} & \beta'_{3,i}\boldsymbol{1}_{|B|}^{\top} \\ \beta'_{2,i}\boldsymbol{1}_{|A|} & \beta'_{2,i}\boldsymbol{1}_{|A|,|A|} + (\beta'_{1,i} - \beta'_{2,i})I_{|A|} & \beta'_{2,i}\boldsymbol{1}_{|A|,|B|} \\ \beta'_{3,i}\boldsymbol{1}_{|B|} & \beta'_{2,i}\boldsymbol{1}_{|B|,|A|} & \beta'_{3,i}\boldsymbol{1}_{|B|,|B|} + (\beta'_{1,i} - \beta'_{3,i})I_{|B|} \end{bmatrix} \quad (18)$$

Here, $R_{0,cont}$ is the basic reproductive number ($R_0$) based on contact. $R_{0,clim}(t)$ is the daily $R_0$ determined by climate conditions using an absolute humidity and temperature-forced model with parameters $R_{0,min,clim}$ and $R_{0,diff,clim}$ (4, 12, 13), and $\overline{R_{0,clim}}$ is its annual mean. Consequently, $R_{0,cont} \frac{R_{0,clim}(t)}{\overline{R_{0,clim}}}$ models seasonal variations of $R_0$ around a mean of $R_{0,cont}$. $\beta'_x$ is the relative transmission rate compared to $\beta_1$ (i.e., $\beta'_1 = 1$). Using relative transmission rates facilitates specifying their initial ranges without requiring specification of their absolute values. $\rho(\boldsymbol{K})$ is the spectral radius of the next generation matrix $\boldsymbol{K}$. $\boldsymbol{N}$ is a diagonal matrix containing population sizes of subpopulations, and $\boldsymbol{\beta}'$ is a matrix containing relative transmission rates $\beta'_x$ within and between subpopulations.

The parts in blue and red represent the traveler network and the migrant worker network, respectively. $f_{trav,i,j}$ is the traveler flow volume from PLAD $j$ to $i$ (i.e., from the local subpopulation $N_j$ to the local subpopulation $N_i$), and $C$ is the set of PLADs other than $i$. $q_{i,j}$ is the proportion of travelers originally from PLAD $i$ among all travelers moving between PLADs $i$ and $j$ (14). $f_{mig,i,j}$ is the migrant worker flow volume from PLAD $j$ to $i$. During the post-CNY period $t_4$, migrant worker flow was separated into $f_{mig,i,j}$ and $f_{mig,i,j,r}$, where $f_{mig,i,j}$ is the volume of new migrant workers from PLAD $j$ to $i$ (i.e., from the local subpopulation $N_j$ in $j$ to the migrant worker subpopulation $N_{i,j}$ in $i$, see Fig. S10e), and $f_{mig,i,j,r}$ is the volume of recurring migrant workers from PLAD $j$ to $i$ (i.e., from the returned migrant worker subpopulation $N_{i,j,r}$ in $j$ to the migrant worker subpopulation $N_{i,j}$ in $i$, see Fig. S10e). Migrant worker flows between host PLAD $i$ and origin PLAD $j$ were modeled as follows (see schematic enclosed by green lines in Figs. 10b–f; note that a host PLAD did not contain a returned migrant worker subpopulation $N_{k,i,r}$):

$$f_{mig,j,i}(t) = \frac{f_{trav,j,i}(t)}{\sum_{t\in t_2} f_{trav,j,i}(t)}(1-\theta)N_{i,j}, t \in t_2 \tag{19}$$

$$f_{mig,i,j}(t) = \frac{f_{trav,i,j}(t)}{\sum_{t\in t_4} f_{trav,i,j}(t)}\left(\frac{N_{i,j,survey}}{\eta} - \left(1-\frac{1}{L}\right)\left(N_{i,j}+N_{i,j,r}\right)\right), t \in t_4 \tag{20}$$

$$f_{mig,i,j,r}(t) = \frac{f_{trav,i,j}(t)}{\sum_{t\in t_4} f_{trav,i,j}(t)}\left(\left(1-\frac{1}{L}\right)\left(N_{i,j}+N_{i,j,r}\right) - N_{i,j}\right), t \in t_4 \tag{21}$$

$$f_{mig,j,i}(t) = \frac{f_{trav,j,i}(t)}{\sum_{t\in t_1'} f_{trav,j,i}(t)}(1-\eta)N_{i,j}, t \in t_1' \tag{22}$$

Here, $\eta$ is the employment rate of migrant workers, $\theta$ is the proportion of migrant workers who remain in host PLADs during CNY, and $L$ is the average duration of stay as migrant workers (see parameter values in Table S3). Specifically, migrant worker flows by time periods were models as follows:

1) During regular period $t_1$ (Fig. 10b), there were no migrant worker flows.

2) During the pre-CNY period $t_2$, to calculate the volumes of returned migrant workers from PLAD $i$ to $j$, $f_{mig,j,i}(t)$, (Eq. 19 and Fig. 10c), we first calculated their total volume as $(1-\theta)N_{i,j}$, and then distributed it proportionally based on daily traveler volumes.

3) During CNY period $t_3$ (Fig.10d), there were no migrant worker flows.

4) During the post-CNY period $t_4$, to calculate the volumes of new migrant workers from PLAD $j$ to $i$, $f_{mig,i,j}(t)$, (Eq. 20 and Fig. 10e), we first calculated the expected total migrant worker population size for the new year, $\frac{N_{i,j,survey}}{\eta}$ (where $N_{survey}$ is the population size from estimated from the national surveys), and the expected population size of migrant workers who would continue working from previous year, $\left(1-\frac{1}{L}\right)\left(N_{i,j}+N_{i,j,r}\right)$. The difference between the two numbers was the total volume of new migrant workers. We then distributed it proportionally based on daily traveler volumes.

5) During period $t_4$, in addition, to calculate the volumes of recurring migrant workers from PLAD $j$ to $i$, $f_{mig,i,j,r}(t)$, (Eq. 21 and Fig. 10e), we first calculated their total volumes by subtracting the population size of migrant workers who remained in PLAD $i$,

$N_{i,j}$, from the expected population size of continuing migrant workers, $\left(1-\frac{1}{L}\right)\left(N_{i,j}+N_{i,j,r}\right)$. We then distributed it proportionally based on daily traveler volumes.

6) During period $t_1'$, to calculate the volumes of migrant workers who failed to secure employment in PLAD $i$ and thus return to $j$ (Eq. 22 and Fig. 10f), we first calculated their total volume as $(1-\eta)N_{i,j}$, and then distributed it proportionally based on the daily traveler volumes.

7) If PLAD $i$ served as an origin PLAD, then the migrant worker flows enclosed by the blue lines in the schematic (Figs. 10b–f; note that an origin PLAD did not contain a migrant worker subpopulation $N_{i,j}$) were modeled using the same framework as described above. If PLAD $i$ served as both a host and an origin PLAD, then the migrant worker flows enclosed by both green and blue lines were combined, exactly as shown in the schematic (Figs. 10b–f). If PLAD $i$ was neither a host nor an origin (note that it only contained the local subpopulation $N_i$), then there were no migrant worker flows throughout the study period.

The numbers of exposed ($E$) and infectious ($I$) individuals seeded through the two networks were drawn from Poisson distributions to simulate the seeding stochasticity.

The parts in green represent the mobility patterns of other migrant groups (such as white-collar migrant workers, who were assumed to have the same immunological profiles as the local subpopulation), to ensure that the simulated total population size of each PLAD aligns with the actual total population size.

**Model initialization.** We estimated the initial range of population susceptibility for each PLAD in 2005 using an age-structured population model. The model simulated susceptibility dynamics across four age groups (0, 1–14, 15–49, and ≥50 years) from 1990 to 2004. The age structure in 1990 was obtained from the national census (15). The simulation started in 1990, when worker migration was limited (16) and regular measles epidemics occurred in most PLADs (17), leading to similar population susceptibilities across PLADs. Thus, we initialized population susceptibility in 1990 using a uniform distribution between 5% and 7% (roughly corresponding to the expected susceptibility for an $R_0$ of 14–20). The model included demographic processes (birth, death, and aging), routine childhood vaccination, and direct transitions from the susceptible ($S$) to recovered/immunized ($R$) compartment due to infection. These infection-driven transitions were based on the estimated incidence by age group. Specifically, incidence by age group was estimated by distributing the annual national incidence from 1990–2004 (18) across PLADs and months based on observed incidence patterns from 2005–2014, and further distributing it among age groups based on distributions reported in the literature (19). The population model accounted for variations in vaccination coverage across PLADs over time, thus allowing its impact on susceptibility to accrue. The resulting population susceptibility for each PLAD $i$ at the end of 2004 was then used as the initial susceptibility in 2005 for both the local subpopulation $N_i$ and the migrant worker subpopulations originating from PLAD $i$, $N_{j,i}$.

Initial ranges of other model state variables for each subpopulation in a PLAD were informed by the initial observed incidence of that PLAD, and model parameters were informed by estimates from literatures (see ranges in Table S4). These state variables include $E$ and $I$, and parameters include $R_{0,cont}$, $R_{0,min,clim}$, $R_{0,diff,clim}$ ($R_{0,min,clim}$ and $R_{0,diff,clim}$ are used to model $R_{0,clim}(t)$ in the absolute humidity and temperature-forced model), $\beta'_x$ ($x\epsilon\{1,2,3\}$), $m_x$ ($x\epsilon\{1,2,3\}$), $Z$, $D$, and $\rho$ (reporting rate, which maps simulated incidence to observed incidence).

**Modeling the nationwide SIA in 2010.** China conducted a nationwide SIA targeting children aged 1–14 years between September 11 and 20, 2010 (20). In the networked metapopulation SEIR model, to model the impact of the SIA on measles epidemic dynamics, individuals were transitioned from the susceptible ($S$) to recovered/immunized ($R$) compartment following the SIA for the local subpopulation, $N_{i,S\to R}$, and the migrant worker subpopulations, $N_{i,j,S\to R}$ (where $j$ indicates an origin PLAD; $N_{sub,S\to R}$ indicates either $N_{i,S\to R}$ or $N_{i,j,S\to R}$), in PLAD $i$ as follows:

$$N_{sub,S\to R} = N_{sub} \times N_{sub,1-14}\% \times \frac{S_{sub,net\ model}\%}{S_{sub,pop\ model}\%} S_{sub,1-14,pop\ model}\% \times \xi_{SIA} \times \frac{1}{L_{SIA}} \quad (23)$$

Here, $N_{sub}$ is the total population size of a subpopulation, and $N_{sub,1-14}\%$ is the proportion of individuals aged 1–14 years in that subpopulation (obtained from the 2010 national census (9) for local subpopulations, and from a migrant worker survey report (21) for migrant worker subpopulations). Because our networked model did not include age structure, we first estimated the susceptibility of the target age group using the aged-structured population model, and then adjust this estimate based on susceptibility from the networked model. Specifically, we ran the population model up to before the SIA to estimate susceptibility for the 1-14-year-old age group, $S_{sub,1-14,pop\ model}\%$, and for the entire subpopulation,

$S_{sub,pop\ model}\%$. We then adjusted $S_{sub,1-14,pop\ model}\%$ using the ratio of susceptibilities of the entire subpopulation from the two models, calculated as $\frac{S_{sub,net\ model}\%}{S_{sub,pop\ model}\%} S_{sub,1-14,pop\ model}\%$. $\xi_{SIA}$ is the SIA effectiveness (defined as the proportion of susceptible individuals in the targeted age group effectively immunized), assumed to be 80% based on the literature (17). $L_{SIA}$ is the duration of the SIA, which was 10 days.

7. Population Census Office under the State Council, Department of Population and Employment Statistics, National Bureau of Statistics of China. 2005 1% National Population Sample Survey. Beijing, China: China Statistics Press; 2005.

8. Population Census Office under the State Council, Department of Population and Employment Statistics, National Bureau of Statistics of China. 2015 1% National Population Sample Survey. Beijing, China: China Statistics Press; 2015.

9. Population Census Office under the State Council, Department of Population and Employment Statistics, National Bureau of Statistics of China. Tabulation on the 2010

Population Census of the People's Republic of China. Beijing, China: China Statistics Press; 2010.

10. National Bureau of Statistics of China. 2025. Available from: https://data.stats.gov.cn/english/.

11. Brauer F, Van den Driessche P, Wu J. Mathematical epidemiology. Berlin, Heidelberg: Springer; 2008.

12. Yuan H, Kramer SC, Lau EHY, Cowling BJ, Yang W. Modeling influenza seasonality in the tropics and subtropics. PLOS Computational Biology. 2021;17(6):e1009050.

13. Iannone R. stationaRy: Detailed meteorological data from stations all over the world. v0.5.1 ed2020.

14. National Tourism Administration of The People's Republic of China. The Yearbook of China Tourism Statistics. Beijing, China: China Travel & Tourism Press; 2015.

15. Population Census Office under the State Council and Department of Population Statistics, State Statistical Bureau of China. Tabulation on the 1990 Population Census of the People's Republic of China. Beijing, China: China Statistical Publishing House; 1990.

16. Lu M, Xia Y. Migration in the People's Republic of China. Tokyo, Japan Asian Development Bank Institute; 2016. Available from: https://www.adb.org/publications/migration-people-republic-china/.

17. Li S, Ma C, Hao L, Su Q, An Z, Ma F, et al. Demographic transition and the dynamics of measles in six provinces in China: A modeling study. PLOS Medicine. 2017;14(4):e1002255.

18. Durrheim DN, Xu A, Baker MG, Hsu LY, Takashima Y. China has the momentum to eliminate measles. The Lancet Regional Health – Western Pacific. 2023;30.

19. Yang W, Li J, Shaman J. Characteristics of measles epidemics in China (1951–2004) and implications for elimination: A case study of three key locations. PLOS Computational Biology. 2019;15(2):e1006806.

20. Ma C, Hao L, Ma J, Zhang Y, Cao L, Liang X, et al. Measles epidemiological characteristics and progress of measles elimination in China, 2010 (in Chinese). Chinese Journal of Vaccines and Immunization. 2011;17(3):242–8.

21. National Bureau of Statistics of China. Nationwide monitoring survey report on rural-to-urban migrant workers (in Chinese) 2014. Available from: https://www.stats.gov.cn/sj/zxfb/202302/t20230203_1898768.html.
